# SleepGPT: A Sleep Stage Language Model for Efficient Sleep Assessment

**DOI:** 10.1101/2024.10.26.24316166

**Authors:** Tianyou Yu, Zhenghui Gu, Zhenfu Wen, Rui Huang, Fei Wang, Man Li, Jingang Yu, Zhuliang Yu, Jun Zhang, Yan Xu, Haiteng Jiang, Wenjuan Liu, Guifeng Deng, Zhengrun Gao, Yiwen Wu, Jun Liu, Yu Zhang, Matt W. Jones, Yuanqing Li, Jun Xiao, Wei Wu

## Abstract

**Background:** Accurate and scalable sleep assessment is essential for the diagnosis of sleep disorders and the advancement of personalized sleep medicine. Current clinical practice relies predominantly on in-laboratory polysomnography (PSG) followed by manual sleep stage scoring. This process is labor-intensive, costly, and difficult to scale to large populations or long-term monitoring scenarios.

**Methods:** We introduce SleepGPT, the first sleep stage language model designed to model the sequential dynamics of sleep macrostructure. SleepGPT was pretrained in a self-supervised manner on over 5.8 million expert-annotated sleep stage labels derived from 5,793 whole-night PSG recordings. The model was evaluated as a plug-in module across multiple state-of-the-art sleep staging architectures and as a sequence-level representation backbone for sleep disorder diagnosis using Transformer-based models. Performance was assessed across multiple publicly available and independent clinical datasets.

**Findings:** Across diverse sleep staging benchmarks (total *N* = 1,320,654 epochs), SleepGPT consistently improved the performance of existing state-of-the-art models and enabled high-accuracy staging from low-density wearable EEG recordings (*N* = 120,095), approaching PSG-level performance. When embedded into a sequence-level Transformer diagnostic framework, SleepGPT accurately identified abnormal sleep stage dynamics and Type-1 narcolepsy across multiple clinical cohorts (total *N* = 685). Attention-based analyses revealed interpretable alterations in sleep macrostructure associated with diagnostic labels.

**Interpretation:** These results demonstrate that SleepGPT functions as a general-purpose foundation model for sleep stage sequence modeling. By capturing the temporal organization of sleep at the macrostructural level, SleepGPT enables scalable, interpretable, and clinically translatable solutions for automated sleep staging and diagnosis of sleep disorders. Notably, operating on sleep stage sequences rather than raw physiological signals may reduce device sensitivity and site-specific variability, supporting more robust deployment across heterogeneous recording settings, including wearable and real-world sleep monitoring.

**Funding:** Details regarding funding supporting this work and all studies involved are provided in the acknowledgements section.

## Introduction

Sleep is essential for health and well-being, with disrupted sleep linked to serious cardiometabolic and neuropsychiatric disorders that collectively affect hundreds of millions worldwide ***Mahowald and Schenck (2005***); ***Zheng et al. (2024***). Comprehensive sleep assessment is foundational to understanding sleep architecture and its contributions to these disorders. Typically, sleep assessment heavily relies on manual scoring of nocturnal polysomnography (PSG) recordings, which is a time-consuming process that requires visually inspecting each 30-second epoch of an entire night’s recording. This results in the generation of a sleep stage sequence, or hypnogram, that characterizes sleep macrostructure, including cycles between rapid eye movement (REM) and non-REM stages, stage durations, transitions, latency, and efficiency. This scoring process is subjective, with inter-scorer reliability often reaching only 82.6% ***Rosenberg and Van Hout (2013***), leading to variability and inefficiencies. Consequently, there is a strong demand for more scalable and objective sleep staging solutions.

Recent advances in machine learning, particularly deep learning, have shown promise for automating sleep staging. Deep neural networks, such as convolutional neural networks (CNNs)***Supratak et al. (2017***); ***Supratak and Guo (2020***), recurrent neural networks (RNNs)***Phan et al. (2019a***,b), transformers***Phan et al. (2022b***), and hybrid networks***Dong et al. (2018***); ***Eldele et al. (2021***); ***Perslev et al. (2021***); ***Phan et al. (2022a***), have been successfully applied to process raw PSG signals or its time-frequency representations to classify sleep stages. However, staging performance often remains suboptimal when contextual information from adjacent epochs was not fully utilized ***Phan et al. (2019a***,b). Sleep staging involves the classification of highly correlated time-series data, where adjacent epochs contain critical contextual information. While recurrent models like long short-term memory (LSTM) networks and gated recurrent units (GRUs) have been employed to address this by capturing long-term dependencies***Phan et al. (2019a***,b); ***Seo et al. (2020***), they still face challenges related to dataset-specific biases and limited generalizability across diverse populations.

In addition to accurate sleep staging, effective sleep assessment should also support the detection and diagnosis of sleep disorders. Traditional diagnostic procedures, such as the multiple sleep latency test (MSLT) for narcolepsy, require significant expertise and are resource-intensive, often leading to diagnostic delays of 7-10 years ***Association (2022***). Moreover, manual analysis by sleep specialists introduces inter- and intra-operator variability ***Thachayani and Loganayagi (2021***), further complicating the diagnostic process. Although automated approaches to sleep disorder diagnosis have been proposed to mitigate these issues, traditional machine learning methods often require extensive feature engineering and suffer from poor generalization to new data. To date, no studies have employed raw sleep stage annotation sequences to diagnose sleep disorders.

Sleep macrostructure is driven by neurophysiological mechanisms that give rise to structured and partially predictable transitions between sleep stages. Leveraging this sequential organization may therefore improve both automated sleep staging and sleep disorder diagnosis ***Phyo et al. (2023***). Inspired by the success of language models in modeling sequential biological data ***Born and Manica (2023***); ***Lin et al. (2023***), we introduce SleepGPT, a sleep stage language model designed to capture the temporal organization of hypnograms. In the sleep staging setting, SleepGPT is used not as a PSG-based classifier, but as a language-model-like sequence prior that refines predictions produced by existing staging models. For sleep disorder diagnosis, the same pretrained model serves as a sequence-level representation learner for hypnogram-based classification. By modeling sleep stage sequences rather than raw physiological signals, SleepGPT provides a general framework for exploiting sleep macrostructure across staging enhancement and diagnostic tasks.

## Methods

### Study design

As illustrated in Fig. 1, SleepGPT was pretrained in a self-supervised manner on millions of expert-annotated sleep stage sequences, learning to predict future sleep stages based on preceding context, analogous to autoregressive language modeling approaches***Radford et al. (2018***, 2019). The pretrained model was subsequently evaluated as a plugin module across multiple downstream sleep-related tasks and datasets.

**Figure 1.**
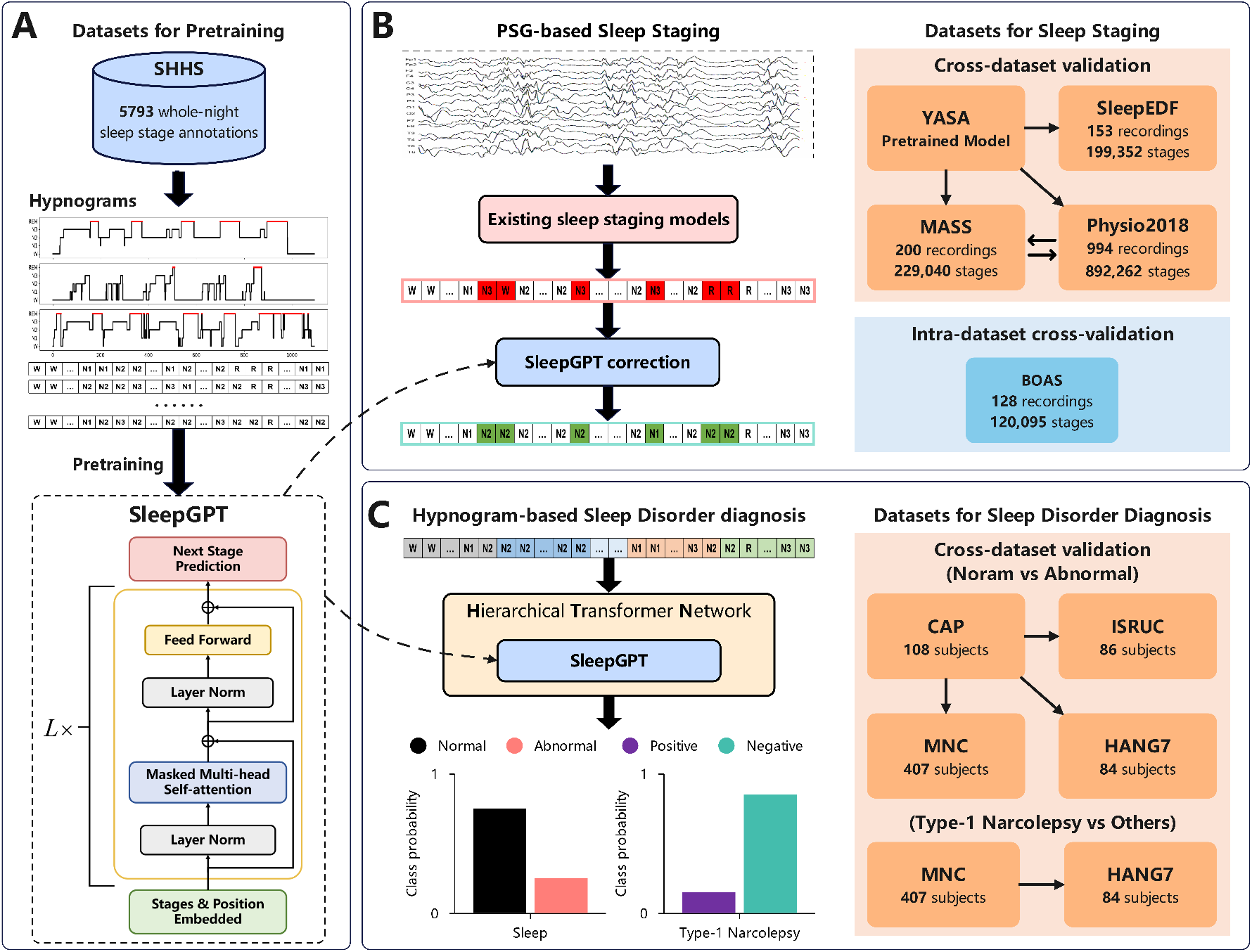
Overview of the proposed SleepGPT framework and its applications to sleep staging enhancement and sleep disorder diagnosis. (**A**) SleepGPT is pretrained on large-scale sleep stage sequences from the SHHS dataset ***Quan et al*. (*1997***), comprising 5.8 million sleep stage annotations derived from 5,793 whole-night recordings. (**B**) Application of SleepGPT to sleep staging enhancement and evaluation protocols. For sleep staging experiments, SleepGPT is evaluated as a post-hoc correction module applied to sleep stage predictions generated by a fixed, publicly available YASA pretrained model. Model generalization is assessed via zero-shot evaluation across multiple independent datasets, including SleepEDF ***Kemp et al. (2000***), MASS ***O’Reilly et al. (2014***), and Physio2018 ***Ghassemi et al. (2018***), without any dataset-specific retraining of the baseline sleep staging model. Moreover, models trained on the MASS dataset are externally validated on Physio2018, and vice versa, to assess cross-dataset generalizability. In addition, the translatability of SleepGPT to wearable EEG settings is evaluated via subject-level cross-validation on the BOAS ***Lopez-Larraz et al. (2024***) dataset, which includes simultaneously acquired PSG and headband EEG recordings. (**C**) Application of SleepGPT to sleep disorder diagnosis. SleepGPT acts as a local sequence-level feature extractor within a hierarchical transformer network (HTN) for hypnogram-based disorder classification. Intra-dataset cross-validation is performed on the CAP ***Terzano et al. (2001***) and MNC ***Stephansen et al. (2018***) datasets. For external validation, models trained on the CAP dataset to distinguish normal from abnormal sleep stage sequences are evaluated on the ISRUC ***Khalighi et al. (2016***), MNC, and HANG7 ***Wang et al. (2023***) datasets, while models trained on the MNC dataset to distinguish Type-1 narcolepsy from other hypersomnia disorders and healthy controls are externally validated on the HANG7 dataset.

For sleep staging, SleepGPT was applied without dataset-specific retraining as a post-hoc sequence refinement module, operating on the outputs of existing staging models. Its impact on performance and generalization was assessed across several independent datasets.

For sleep disorder diagnosis, SleepGPT was embedded as a local sequence feature extractor within a hierarchical Transformer network (HTN), enabling joint modeling of local sleep-stage transitions and global whole-night sleep architecture for classification. Diagnostic performance was evaluated using intra-dataset cross-validation and external validation on previously unseen cohorts.

### Cohorts and ethics

This study involves eight publicly available sleep datasets and one private sleep dataset (detailed information on these data sets is described in Supplementary Materials p2-3):

#### Pretraining datasets

The Sleep Heart Health Study (SHHS) is a multi-center cohort examining the cardiovascular and other consequences of sleep-disordered breathing ***Quan et al. (1997***); ***Zhang et al. (2018***).

#### Sleep staging datasets

The SleepEDF Expanded dataset is a collection of overnight PSG recordings from 78 healthy Caucasian subjects aged 25 to 101 years ***Kemp et al. (2000***); ***Goldberger et al. (2000***). The Montreal Archive of Sleep Studies (MASS) database comprises whole-night hospital sleep laboratory recordings from 200 subjects (97 males and 103 females) aged 18 to 76 years ***O’Reilly et al. (2014***). The PhysioNet 2018 Challenge dataset comprises 1,985 polysomnographic recordings provided by the Computational Clinical Neurophysiology Laboratory (CCNL) and the Clinical Data Animation Center (CDAC) at Massachusetts General Hospital (MGH) ***Ghassemi et al. (2018***); ***Goldberger et al. (2000***). The Bitbrain Open Access Sleep (BOAS) dataset serves to bridge the gap between gold-standard clinical sleep monitoring and emerging wearable EEG technologies ***Lopez-Larraz et al. (2024***).

#### Sleep disorder diagnosis datasets

The Cyclic Alternating Pattern (CAP) Sleep Database is a collection of 108 polysomnographic recordings contributed by the Sleep Disorders Center of the Ospedale Maggiore of Parma, Italy ***Terzano et al. (2001***); ***Goldberger et al. (2000***) (Table **S3**). The ISRUC-Sleep dataset contains full-night PSG recordings collected at the Sleep Medicine Centre of Coimbra University Hospital (CHUC) between 2009 and 2013 ***Khalighi et al. (2016***). The Mignot Nature Communications (MNC) dataset comprises raw polysomnography data collected from an automated sleep staging project utilizing neural networks ***Stephansen et al. (2018***); ***Zhang et al. (2018***) (Table **S4**). The HANG7 dataset collected 84 participants aged 11 to 57 years polysomnography recordings at the Affiliated Mental Health Center & Hangzhou Seventh People’s Hospital, Zhejiang University School of Medicine ***Wang et al. (2023***). This study was conducted in accordance with the Declaration of Helsinki and approved by the Institutional Review Board of the Ethics Committee of Hangzhou Seventh People’s Hospital, China (Approval No. 2023-053-02). Written informed consent was obtained from all participants or their legal caregivers prior to participation.

### Model development

SleepGPT is developed based on the GPT-2 architecture ***Radford et al. (2019***), which is designed to capture long-range dependencies in sequential data. The model is pretrained exclusively on the SHHS-1 dataset ***Quan et al. (1997***), comprising approximately 5.8 million sleep stage annotations from 5,793 subjects with whole-night PSG recordings. Each subject’s recording is represented as a sequence of 30-second sleep stage annotations, ranging from 360 to 1,199 epochs per night (mean = 1,012), yielding a total of 5,863,207 labeled epochs.

Pretraining is formulated as a next-stage prediction task, in which SleepGPT learns to predict the subsequent sleep stage given a sequence of preceding stages. This objective encourages the model to learn latent temporal patterns, transition regularities, and long-range dependencies characteristic of human sleep macrostructure. The pretrained SleepGPT model is fixed and reused across all downstream experiments.

### Evaluation protocols: sleep staging enhancement

SleepGPT is evaluated as a post-hoc sequence-context correction module for predictions generated by existing sleep staging models. Importantly, SleepGPT is not a standalone staging model that maps PSG signals directly to sleep-stage labels. Rather, it operates on sleep-stage tokens after an initial PSG-based prediction has been made. In this framework, the staging model provides signal-driven evidence for each epoch, whereas SleepGPT provides a learned prior over physiologically plausible sleep-stage sequences based on expert-scored hypnograms. This division of labor is analogous to automatic speech recognition, in which an acoustic model provides signal-based evidence and a language model improves decoding by modeling learned sequential regularities. Sleep staging performance is assessed on three publicly available PSG datasets: SleepEDF ***Kemp et al. (2000***), MASS ***O’Reilly et al. (2014***), and Physio2018 ***Ghassemi et al. (2018***). In all experiments, SleepGPT is applied only at inference time, without retraining the baseline staging models, and performance with and without SleepGPT is compared to quantify its contribution.

Three complementary evaluation protocols are designed to comprehensively assess the effectiveness of SleepGPT. First, to assess zero-shot generalization, we employ the YASA toolbox ***Vallat and Walker (2021***), which provides pretrained, publicly available sleep staging models. In this setting, all YASA model parameters are fixed prior to evaluation, and SleepGPT is used as an add-on correction module across SleepEDF, MASS, and Physio2018, enabling blinded out-of-sample evaluation.

Second, inter-cohort generalization is examined by training sleep staging models on one dataset (e.g., MASS) and evaluating them on another unseen dataset (e.g., Physio2018), and vice versa. SleepGPT is applied consistently during inference without modifying the training procedure of the base models.

Third, to examine translational potential in wearable settings, SleepGPT is further evaluated on the BOAS dataset ***Lopez-Larraz et al. (2024***), which includes simultaneous PSG and low-density wearable EEG recordings from 128 subjects. Subject-level cross-validation is performed to compare sleep staging performance across modalities, with and without SleepGPT integration.

### Evaluation protocols: sleep disorder diagnosis

For hypnogram-based sleep disorder diagnosis, SleepGPT is incorporated as a local sequence-level feature extractor within a hierarchical transformer network (HTN) (Supplementary Fig. **S1**). This architecture captures both local sleep stage patterns and global sequence-level representations.

Intra-dataset subject-level cross-validation is conducted on the CAP dataset ***Terzano et al. (2001***) to distinguish normal from abnormal sleep stage sequences. To assess generalizability, models trained on the full CAP dataset are externally evaluated on three independent datasets: ISRUC ***Khalighi et al. (2016***), MNC ***Stephansen et al. (2018***), and HANG7 ***Wang et al. (2023***).

For a more clinically challenging task, SleepGPT-based models are evaluated for distinguishing Type-1 narcolepsy from other hypersomnia disorders and healthy controls. This analysis is performed using subject-level cross-validation on the MNC dataset, followed by external validation on the HANG7 dataset. In addition, global attention weights learned by the HTN model are visualized to identify salient sequence-level patterns that may serve as candidate biomarkers.

## Results

We begin by assessing whether SleepGPT can consistently enhance sleep staging performance across multiple datasets and evaluation settings.

### Enhancing sleep staging with SleepGPT

We evaluated SleepGPT’s ability to enhance automated sleep staging by post-hoc correction of predictions made by established models, including TinySleepNet ***Supratak and Guo (2020***) and XSleepNet ***Phan et al. (2022a***). Performance was assessed in terms of accuracy, macro-averaged F1-score (MF1), and Cohen’s kappa coefficient (*κ*) across multiple external datasets. Note that SleepGPT was pretrained on the SHHS-1 dataset and was not fine-tuned on any of the downstream datasets used in this evaluation. To assess generalization, we applied several state-of-the-art sleep staging models trained on one dataset to unseen data from another, and tested whether SleepGPT could improve their out-of-sample predictions. Specifically, sleep staging models (TinySleepNet and XSleepNet) trained on the MASS dataset (N=229,040) were tested on the Physio2018 dataset (N=892,262), and vice versa, enabling cross-dataset validation with matching EEG configurations (C3-A2/C4-A1). We also applied SleepGPT to enhance predictions made by YASA (Yet Another Spindle Algorithm) ***Vallat and Walker (2021***), a widely used open-source sleep staging toolbox with fixed pretrained models, on the SleepEDF and MASS datasets.

As shown in Fig. 2, integrating SleepGPT consistently improved the performance of all baseline models across all datasets and model architectures. YASA’s accuracy improved by 4.2% on SleepEDF and 1.6% on MASS. TinySleepNet’s accuracy increased by 2.9% and 1.6%, and XSleepNet’s by 2.3% and 1.9%, during cross-dataset validation between the Physio2018 and MASS datasets. Consistent improvements in MF1, *κ*, and stage-specific accuracies are also observed. Notably, models trained on the larger Physio2018 dataset (N=892,262) and tested on the smaller MASS dataset (N=229,040) demonstrated superior staging performance than the reverse, suggesting that the size and composition of the training dataset, particularly the distribution of sleep stages, have a considerable impact on model generalizability. Examples of the corrections made by SleepGPT during cross-dataset validation are shown in Fig. 2F (more detailed results are provided in Table **S5** of the Supplementary Appendix), illustrating the ability of SleepGPT in correcting the erroneous sleep stage predictions made by baseline models. Overall, these results demonstrate SleepGPT’s robustness and utility as a general-purpose add-on module that improves sleep staging performance across diverse datasets and model architectures.

**Figure 2.**
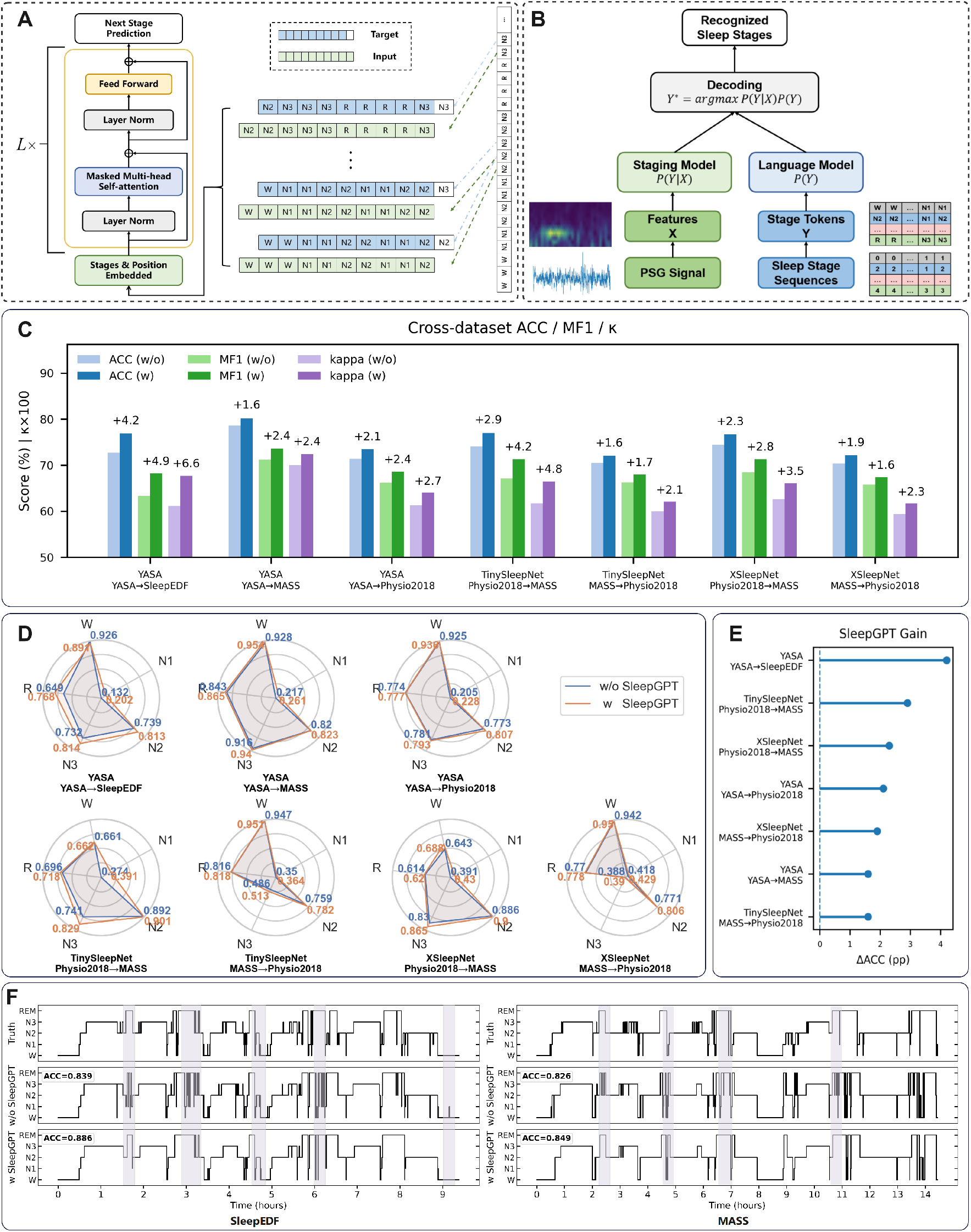
Results of three state-of-the-art staging methods with and without SleepGPT when performing cross-dataset sleep staging on the SleepEDF, MASS, and Physio2018 datasets. The labels **Source-to-Target** indicate that the staging model is trained from the **Source** dataset and evaluated on the **Target** dataset. (**A**) Architecture and training procedure of the SleepGPT model. (**B**) The pipeline for enhancing existing sleep staging models with SleepGPT. (**C-E**) Performance comparison of YASA ***Vallat and Walker (2021***), TinySleepNet ***Supratak and Guo (2020***), and XSleepNet ***Phan et al. (2022a***) with and without SleepGPT on the SleepEDF, MASS, and Physio2018 datasets. (**F**) Two examples of the corrections made by SleepGPT during cross-dataset validation. The shaded areas indicate where the SleepGPT model corrected the erroneous sleep stage predictions of the baseline staging models.

### Assessing translatability of SleepGPT-powered models on wearable EEG data

Recent advances in wearable EEG devices have created new opportunities for low-burden, at-home sleep monitoring. To assess the translational potential of SleepGPT, we evaluated its ability to enhance sleep staging performance using data from the BOAS dataset, which contains simultaneous PSG and headband EEG recordings from 128 subjects. The TinySleepNet ***Supratak and Guo (2020***) and XSleepNet ***Phan et al. (2022a***) models, with and without SleepGPT integration, were cross-validated on the PSG and the headband EEG data, respectively. As shown in Fig. 3, both models exhibit comparable performance across the PSG and headband EEG data, and the addition of SleepGPT consistently improved accuracy, MF1, and *κ* scores across both modalities. Notably, the sleepGPT-powered XSleepNet model achieved the highest accuracy of 84.3% and 83.9% for the PSG and headband EEG data, respectively. More detailed results are provided in Table **S6** of the Supplementary Appendix. These results underscore the robustness of the SleepGPT models in enhancing sleep staging performance across EEG modalities and support their potential for integration into wearable sleep monitoring applications, enabling adoption of low-burden, low-cost, longitudinal sleep monitoring without significantly compromising accuracy.

**Figure 3.**
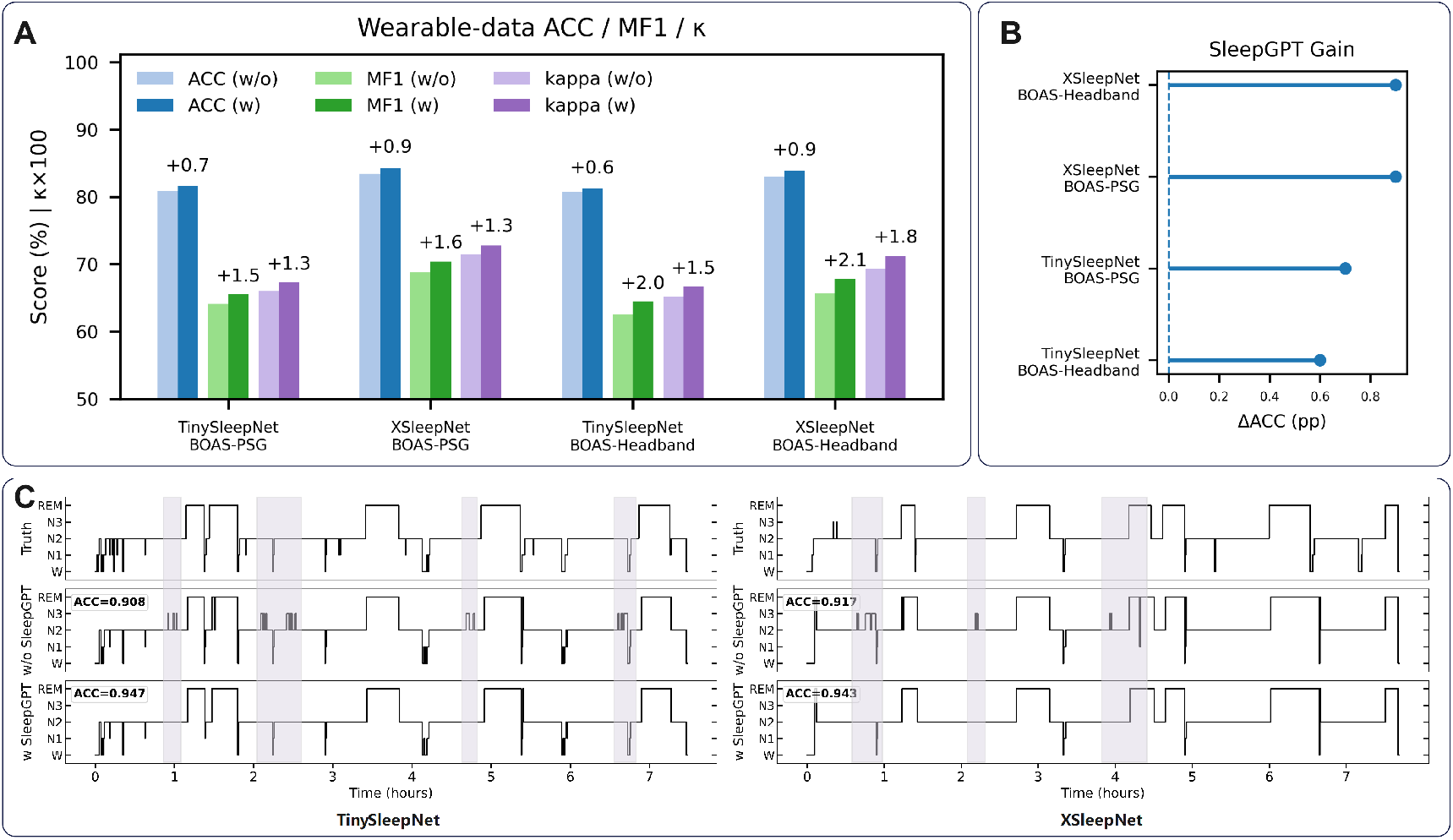
Sleep staging results of the state-of-the-art staging methods with and without SleepGPT on the BOAS wearable dataset. (**A**) Performance comparison of TinySleepNet ***Supratak and Guo (2020***) and XSleepNet ***Phan et al.(2022a***) with and without SleepGPT on the PSG and headband EEG data. (**B**) Performance gain obtained by integrating SleepGPT into the TinySleepNet and XSleepNet models on the PSG and headband EEG data. (**C**) Examples of the corrections made by SleepGPT on the headband EEG data. The shaded areas indicate where the SleepGPT model corrected the erroneous sleep stage predictions of the baseline staging models.

### Sleep disorder diagnosis enhanced by SleepGPT

We next evaluated SleepGPT as a feature extractor for sleep disorder diagnosis, comparing its performance to state-of-the-art diagnostic methods. Specifically, we integrated SleepGPT into a hierarchical transformer network (HTN) that takes a subject’s full-night sleep stage sequence as input, enabling it to leverage the temporal dynamics of sleep stage transitions for disorder identification. Receiver operating characteristic (ROC) curves and confusion matrices were used to assess diagnostic performance across the modified CAP ***Terzano et al. (2001***), ISRUC ***Khalighi et al. (2016***), MNC ***Stephansen et al. (2018***), and HANG7 datasets. Fig. 4 presents results for two empirical feature-based XGBoost classifiers (based on Hypnogram ***Vallat and Walker (2021***) and Hypnodensity ***Stephansen et al. (2018***)), a baseline end-to-end neural network (BaseNet), the proposed HTN trained from scratch, and the HTN model initialized with pretrained SleepGPT parameters. ROC curves and confusion matrices for each method across the four datasets are shown in Fig. 4A and 4B, with detailed results provided in Table **S7** and **S8**.

**Figure 4.**
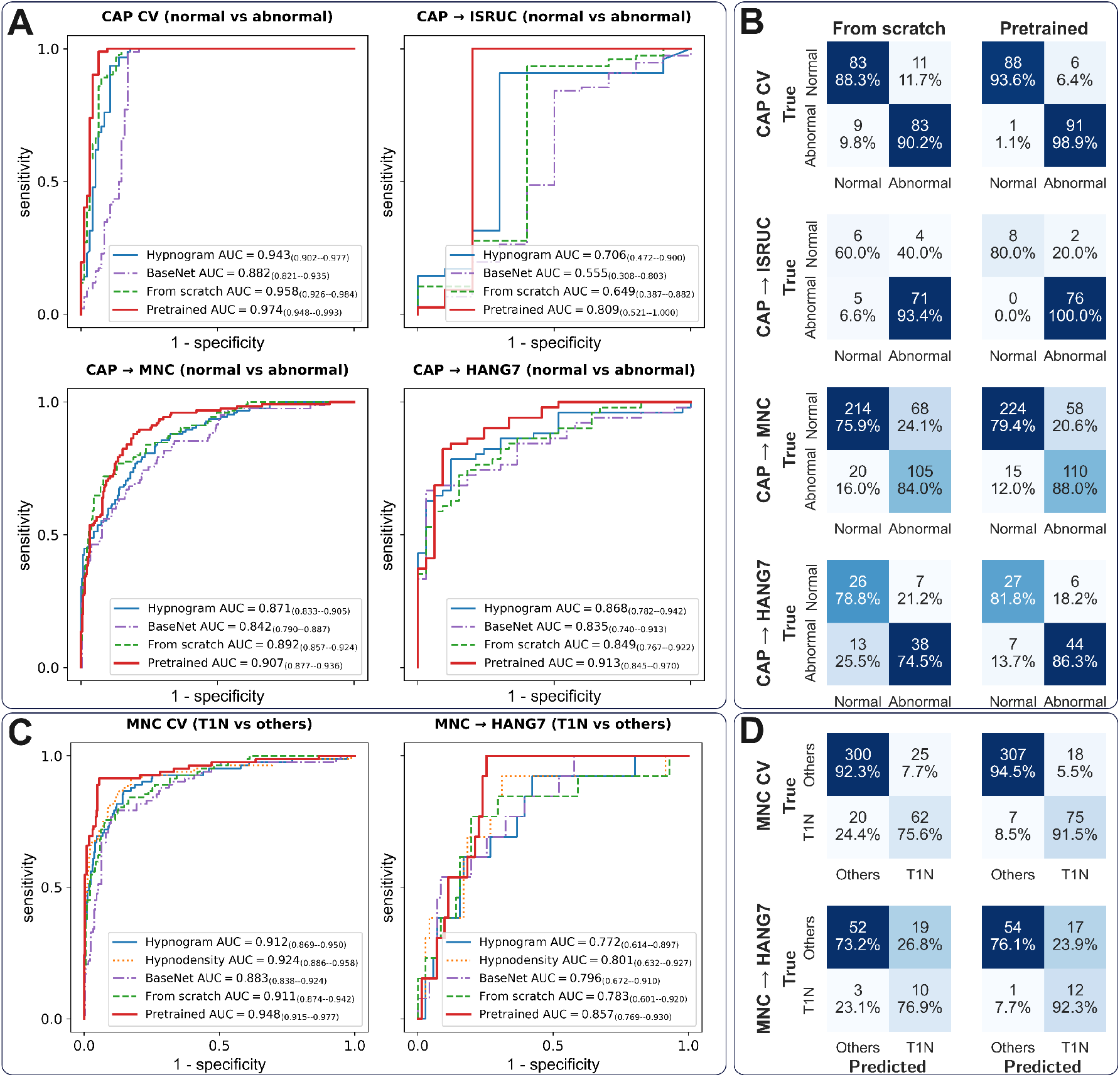
Performance of the SleepGPT-powered hierarchical transformer network (HTN) and baseline methods for sleep disorder diagnosis, including normal vs abnormal sleep classification on the CAP, ISRUC, MNC, and HANG7 datasets, and Type-1 narcolepsy (T1N) vs non-T1N classification on the MNC and HANG7 datasets. (**A**) Receiver operating characteristic (ROC) curves for abnormal sleep detection. Cross-validation (CV) results on CAP and external validation results on ISRUC, MNC, and HANG7 are shown. The area under the ROC curve (AUC) is reported in the legend together with the corresponding 95% confidence intervals (CI), estimated using bootstrap resampling. (**B**) Confusion matrices for HTN models trained from scratch and with pretrained SleepGPT parameters across the four datasets. (**C**) ROC curves for T1N detection, showing CV results on MNC and external validation on HANG7. AUC values with 95% CI are reported in the legend. (**D**) Confusion matrices for HTN models trained from scratch and with pretrained SleepGPT parameters on both datasets.

On the CAP dataset, in which the task was to distinguish between normal sleep and abnormal sleep (including sleep disorders such as insomnia, disordered breathing, narcolepsy, etc.), the XG-Boost classifier trained with hypnogram features achieved a balanced accuracy of 90.91%, with a sensitivity of 95.65% and a specificity of 86.17%. While end-to-end models like BaseNet can learn useful features from hypnograms, they may struggle to capture the sequential attributes of sleep stages if the model architecture is not carefully designed, as evidenced by a lower balanced accuracy of 84.97%, sensitivity of 86.96%, and specificity of 82.98%. The SleepGPT-based HTN model, however, significantly improved performance in an end-to-end fashion. This highlights the HTN’s capacity to capture both localized sequential sleep transitions and broader contextual patterns in sleep hypnograms, enhancing diagnostic performance. Notably, initializing the HTN with pre-trained SleepGPT parameters yielded the highest performance, achieving a balanced accuracy of 96.27%, with a sensitivity of 98.91% and a specificity of 93.62%. This pretrained HTN demonstrated a marked improvement in sensitivity over its non-pretrained counterpart, with an increase of over 10% on the CAP dataset.

Similar trends were observed in the MNC dataset, which involved distinguishing Type-1 narcolepsy (T1N) patients from non-T1N individuals (including other hypersomnia patients and healthy controls), an arguably more challenging task than distinguishing general sleep-disorder patients from healthy controls. The XGBoost classifier trained on hypnogram and hypnodensity features achieved balanced accuracies of 84.39% and 85.16%, respectively. Although the BaseNet model performed reasonably well in diagnosing T1N patients, achieving a balanced accuracy of 79.69%, it struggled with sensitivity, identifying only 67.07% of T1N subjects. The HTN model trained from scratch achieved balanced accuracy of 85.49%, sensitivity of 78.05%, and specificity of 92.92%, matching the performance of the XGBoost classifier trained on hypnodensity features. When fine-tuned with pretrained SleepGPT parameters, the HTN model further boosted performance, achieving a balanced accuracy of 92.81%, a sensitivity of 91.46%, and a specificity of 94.15%. This improvement underscores the benefits of the SleepGPT model’s self-supervised pretraining on large-scale sleep stage datasets. Importantly, the pretrained SleepGPT model demonstrated the potential for fine-tuning on smaller datasets, which is crucial in sleep medicine, where data scarcity often presents a challenge.

### Testing generalization of SleepGPT-powered diagnosis models

We further assessed the out-of-sample generalizability of SleepGPT-powered models in diagnosing abnormal sleep. We evaluated the prediction performance by training models using the CAP dataset, and directly applied them to the ISRUC, MNC, and HANG7 datasets. The ISRUC sleep dataset contains whole-night PSG recordings of obstructive sleep apnea (OSA) patients (N=76) and healthy controls (N=10). On this external dataset, the pretrained HTN model achieved a balanced accuracy of 90.00%, with a sensitivity of 100% and a specificity of 80.00% when distinguishing OSA patients from healthy controls (Fig. 4 and Table **S7**). When applied to the MNC dataset, the same model achieved a balanced accuracy of 83.72%, with a sensitivity of 88.00% and a specificity of 79.43% in discriminating healthy controls (N=282) from patients with T1N or other hypersomnia (N=125). On the HANG7 dataset, which contains 51 patients with narcolepsy and 33 healthy controls, the model achieved a balanced accuracy of 84.05%, with a sensitivity of 86.27% and a specificity of 81.82%.

For the more challenging task of differentiating T1N patients from other hypersomnia patients and healthy controls, the HTN model trained on the MNC dataset was externally validated using the HANG7 dataset. As shown in Fig. 5 (and in Table **S8**), the HTN model fine-tuned with SleepGPT parameters achieved a balanced accuracy of 84.18%, with a sensitivity of 92.31% and a specificity of 76.06%, outperforming the compared methods. In all settings, SleepGPT-based models outperformed conventional baselines, demonstrating robust generalization across datasets and diagnostic categories.

**Figure 5.**
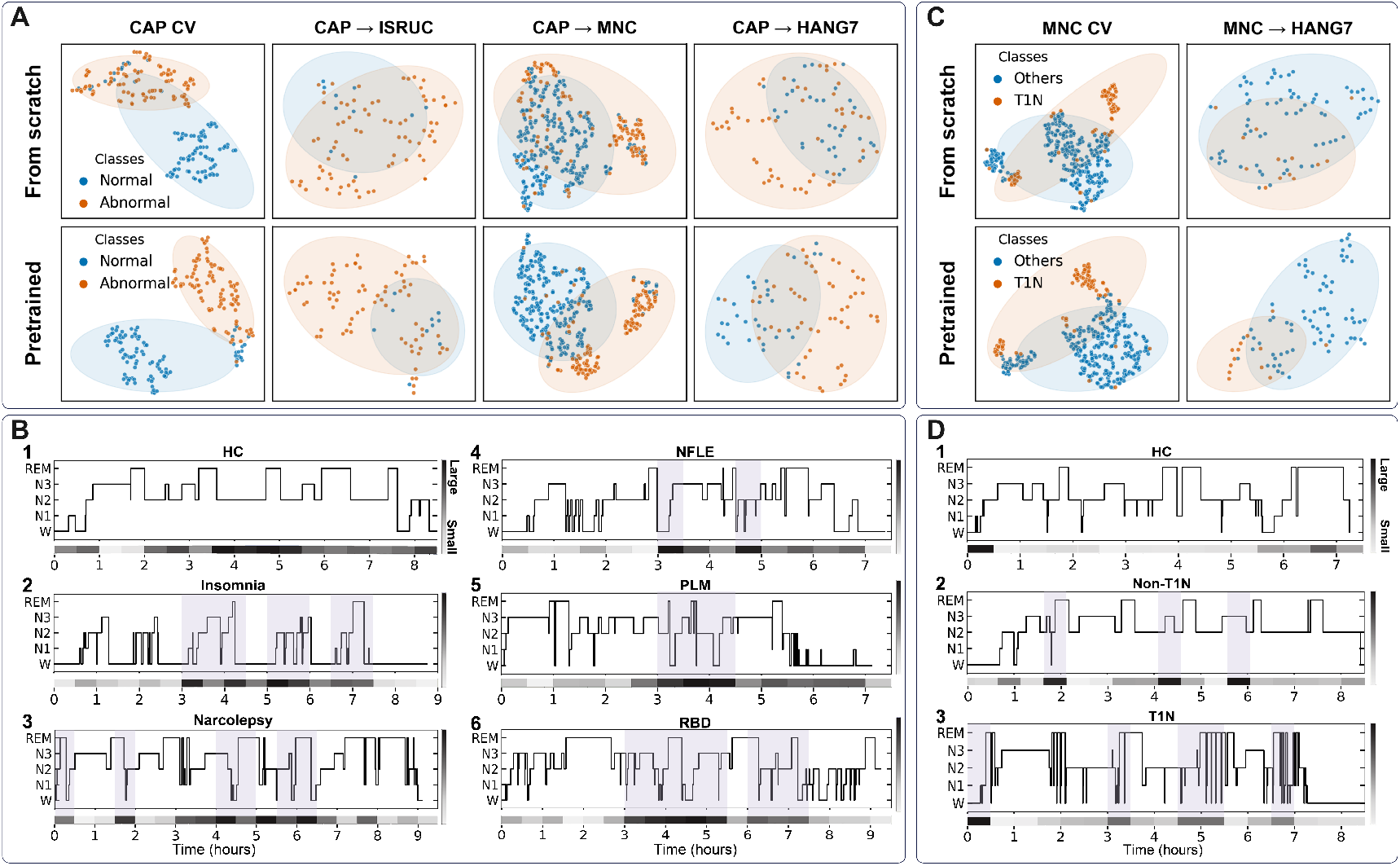
Visualization and interpretation of the SleepGPT-powered hierarchical transformer network (HTN) for sleep disorder diagnosis. (**A**) t-SNE visualization of features extracted by HTN with and without SleepGPT pretraining on CAP dataset. (**B**) Visualization of global transformer encoder attention weights on CAP, highlighting healthy and representative subjects with insomnia, narcolepsy, NFLE, PLMs, and RBD. Gray-shaded areas indicate segments with abnormal sleep patterns. (**C**) t-SNE visualization of features extracted by HTN with and without SleepGPT pretraining on MNC dataset. (**D**) Visualization of global transformer encoder attention weights on MNC, highlighting representative subjects (healthy, T1N, other hypersomnia); gray-shaded areas indicate segments with discriminative sleep patterns.

### Interpretation of the SleepGPT-powered diagnosis models

To better understand feature separability, t-SNE visualizations of the SleepGPT-powered HTN model for sleep disorder diagnosis are provided in Fig. 5A and Fig. 5C. The features extracted by the HTN model with pretrained SleepGPT parameters show a more distinct separation between normal/T1N and abnormal/non-T1N sleep patterns compared to the features extracted by the HTN model trained from scratch. This indicates that the pretrained SleepGPT model effectively captures discriminative features relevant to sleep disorder diagnosis, enhancing the model’s ability to differentiate between normal/T1N and abnormal/non-T1N sleep patterns.

Moreover, to provide insights into the global feature extractor of the HTN model, a visualization of the attention weights from the global feature extractor on the CAP dataset is presented in Fig. 5B for one healthy subject and five subjects with distinct sleep disorders—insomnia, narcolepsy, nocturnal frontal lobe epilepsy (NFLE), periodic leg movements (PLMs), and REM behavior disorder (RBD). By focusing on the attention weights assigned to the *CLS* token (a special token that outputs a global representation of the entire sequence, detailed in the Methods section) within the final transformer layer, we can observe how each model identifies and prioritizes salient features in the data. The results demonstrate the effectiveness of the global feature extractor in capturing atypical sleep patterns associated with sleep disorders. For instance, in the case of the insomnia subject, characterized by small N3 and REM sleep ratios and direct transitions from REM sleep to wakefulness, the global feature extractor allocates more attention to the segments marked by these transitions (Fig. 5B(2)). Likewise, the global transformer encoder of HTN highlights segments with short wake-to-REM sleep latency or abrupt transitions from wakefulness to REM sleep for the narcolepsy subject (Fig. 5B(3)). Furthermore, the self-attention module identifies segments featuring abnormal stage transitions, short N3 and REM sleep durations, direct shifts from REM sleep to wakefulness, and frequent toggling between deep (N3) or REM sleep and wakefulness (Fig. 5B(4-6)). Conversely, for subjects without pathologies, the attention weights are more evenly dispersed across the hypnogram segments (Fig. 5B(1)).

We also show the attention weights obtained by the global feature extractor of the HTN model on the MNC data set for a healthy control, a patient with hypersomnia, and a patient with T1N (Fig. 5D). For the healthy control, attention weights are more evenly distributed across the hypnogram segments. While for the narcolepsy patient, there is increased attention to segments featuring short wake-to-REM latency or direct transitions from wakefulness to REM sleep. Since we classify T1N patients from other hypersomnia patients and healthy controls, the attention weights are more focused on segments similar to those of the narcolepsy patient. These segments include short wake-to-REM latency, direct transitions from wakefulness to REM sleep, and dissociated REM sleep. These results suggest that the HTN’s attention mechanism captures meaningful macrostructural patterns associated with specific sleep disorders. This interpretability not only enhances model trustworthiness but also points to potential biomarkers for clinical diagnosis.

## Discussion

This study introduces SleepGPT, a novel sleep language model adapted from the GPT architecture and trained in a self-supervised manner on a comprehensive sleep stage dataset. SleepGPT offers several advantages for efficient sleep assessment: it consistently enhances existing sleep staging methods, effectively captures sleep stage transition dynamics, and integrates as a feature extractor within hierarchical transformer networks to improve sleep disorder diagnosis. Additionally, it identifies interpretable abnormal sleep patterns, potentially providing mechanistic insights into sleep disorders. Taken together, these findings highlight SleepGPT’s potential as a scalable, clinically translatable artificial intelligence (AI)-powered solution for automated sleep assessment.

In sleep staging, SleepGPT’s attention mechanisms focus on critical stage transitions, identifying inconsistent patterns, such as abrupt shifts from deep sleep (N3) directly to wakefulness, that may signal staging errors. The benefits of SleepGPT are particularly pronounced in blinded evaluations on independent datasets, where data heterogeneity often hinders the generalizability of sleep staging models. By effectively capturing the intrinsic sequential patterns of sleep stages, SleepGPT enhances model generalizability across diverse datasets.

In contrast to traditional sleep disorder diagnosis approaches that rely on handcrafted features derived from hypnogram analysis (e.g., stage latencies, durations, and transitions), our proposed HTN directly models and categorizes sleep stage sequences in an end-to-end fashion. This hierarchical architecture allows the model to capture both local and global contextual information within sleep hypnograms, enabling more accurate classification of sleep stage sequences, especially those with varying lengths, compared to the baseline model. Notably, the HTN effectively addresses the challenge of subject-level sleep disorder diagnosis in weakly supervised learning scenarios, where only session-level labels are available and sleep disorder symptoms may not be consistently present throughout the entire sleep session ***Xu et al. (2022***). As such, the HTN achieves accurate subject-level classification and identifies potential biomarkers indicative of specific sleep disorders by leveraging the self-attention mechanism in its global transformer module. These insights hold promise for developing novel diagnostic methodologies and designing precision therapies. Moreover, the HTN’s adaptability allows for its potential application to other sleep-related brain disorders, such as depression, anxiety, dementia, and Parkinson’s disease ***Mahowald and Schenck (2005***); ***Zheng et al. (2024***), e.g., by replacing sleep disorder labels with corresponding condition labels. This flexibility is particularly valuable given the frequent comorbidity of sleep disorders with other neurological or psychiatric conditions. Additionally, operating on sleep stage sequences rather than raw physiological signals may reduce sensitivity to device- and site-specific variability, supporting more robust deployment across heterogeneous recording settings, including wearable and real-world sleep monitoring.

On the CAP dataset, the pretrained HTN model effectively identifies nearly all abnormal subjects, with only 1.09% of abnormal subjects being erroneously classified as normal. This high sensitivity is crucial in sleep disorder diagnosis, where false positives (misclassifying normal as abnormal) are generally preferable to false negatives, as they lead to further medical investigation, while undetected abnormalities could have serious consequences. Moreover, previous literature indicates that sensitivity and specificity for T1N are 75-90% and 90-98%, respectively ***Stephansen et al. (2018***). The performance achieved by the pretrained HTN model on the MNC dataset is comparable to or exceeds these reported values. These results suggest that analyzing a single-night PSG can be as effective as the PSG-MSLT gold standard, which involves a 24-hour procedure and is expensive. Consequently, our model offers a reliable and cost-effective alternative as a screening tool for T1N, streamlining the pathway for patients requiring further diagnostic evaluation.

The strong performance of SleepGPT-based models in sleep staging with wearable EEG and disorder identification opens exciting possibilities for real-time, at-home sleep monitoring. Wearable devices powered by SleepGPT could provide immediate feedback on sleep quality, identify potential sleep disturbances as they occur, and offer personalized advice for improving sleep. Such real-time monitoring could be invaluable for individuals with chronic sleep disorders, allowing for timely interventions and adjustments to treatment plans. Furthermore, continuous sleep monitoring could facilitate early detection of sleep problems, potentially preventing them from escalating into more serious health issues.

While our experiments demonstrate the effectiveness of SleepGPT, it is important to acknowledge certain limitations. The SHHS dataset used for pretraining SleepGPT is relatively small compared to the massive text corpora used to train traditional language models. Expanding the pretraining dataset could further enhance model performance. Future research should also explore fine-grained sleep disorder classification by leveraging larger and more diverse datasets to gain deeper insights into specific sleep stage patterns and transitions associated with particular disorders. Furthermore, while hypnograms provide valuable information on sleep macrostructure, they have limited ability to capture microstructural events like sleep spindles and K-complexes. Integrating additional data modalities, such as ECG or other physiological signals, may enhance the accuracy and granularity of sleep disorder diagnosis. Finally, the present study is retrospective and based on curated sleep datasets, which may introduce dataset-specific biases and limit generalizability. Although multiple independent cohorts were included to evaluate cross-dataset performance, the datasets were collected under specific clinical protocols and inclusion criteria. As a result, distributional differences across populations and recording settings may affect model behavior. In addition, the proposed framework has not yet been deployed or evaluated in prospective clinical workflows and should be viewed as a decision-support and screening tool rather than a standalone diagnostic system. Prospective validation in real-world clinical environments will therefore be necessary to fully assess robustness and clinical utility.

## Conclusion

In summary, this study presents SleepGPT, a novel GPT-based sleep language model designed to advance automated sleep staging and sleep disorder diagnosis. Through extensive evaluation across multiple publicly available and independent datasets, SleepGPT demonstrated consistent improvements in sleep staging accuracy and generalizability, outperforming existing state-of-the-art models. Its integration as a feature extractor within hierarchical transformer networks further enabled robust and scalable sleep disorder classification, including challenging tasks such as Type-1 narcolepsy identification. The model’s ability to capture sequential sleep stage dynamics and highlight interpretable patterns underscores its potential for clinical translation and precision sleep medicine. Future work may focus on expanding pretraining datasets, refining model architectures, and integrating multimodal physiological signals to further enhance diagnostic capabilities and applicability in real-world settings.

## Data Availability

The SHHS and MNC datasets are provided by the National Sleep Research Resource with appropriate deidentification. Permission and access for these datasets can be obtained via the online portal:
https://www.sleepdata.org. The SleepEDF, Physio2018, and CAP datasets are available from PhysioNet at https://physionet.org/content/sleep-edfx/1.0.0/, https://physionet.org/content/challenge-2018/1.0.0/ and https://physionet.org/content/capslpdb/1.0.0/, respectively. The MASS dataset is available at http://ceams-carsm.ca/mass/. The BOAS dataset can be accessed at https://openneuro.org/datasets/ds005555/versions/1.0.0. The ISRUC dataset can be accessed at https://sleeptight.isr.uc.pt/}. Access to the HANG7 dataset is governed by data-use agreements, and it is therefore not publicly available.

## Contributors

T.Y. contributed to the development of methods, the analysis and interpretation of the data, and the drafting of the manuscript. Z.G. contributed to the development of methods, the analysis and interpretation of the data, and the drafting of the manuscript. Z.W., R.H., F.W., M.L., J.Y., Z.Y., J.Z., Y.X., H.J., W.L., G.D., Z.G., Y.W., J.L., Y.Z., and M.J. contributed to the analysis and interpretation of the data. Y.L. contributed to the analysis and interpretation of the data and the drafting of the manuscript. J.X. contributed to the development of methods, the analysis and interpretation of the data, and the drafting of the manuscript. W.W. contributed to the development of methods, the analysis and interpretation of the data, and the drafting of the manuscript. All authors reviewed the manuscript.

## Data sharing statement

The SHHS ***Quan et al. (1997***) and MNC ***Stephansen et al. (2018***) datasets are provided by the National Sleep Research Resource with appropriate deidentification. Permission and access for these datasets can be obtained via the online portal: https://www.sleepdata.org. The SleepEDF ***Kemp et al. (2000***), Physio2018 ***Ghassemi et al. (2018***), and CAP ***Terzano et al. (2001***) datasets are available from PhysioNet at https://physionet.org/content/sleep-edfx/1.0.0/, https://physionet.org/content/challenge2018/1.0.0/, and https://physionet.org/content/capslpdb/1.0.0/, respectively. The MASS ***O’Reilly et al. (2014***) dataset is available at http://ceams-carsm.ca/mass/. The BOAS ***Lopez-Larraz et al. (2024***) dataset can be accessed at https://openneuro.org/datasets/ds005555/versions/1.0.0. The ISRUC ***Khalighi et al. (2016***) dataset can be accessed at https://sleeptight.isr.uc.pt/. Access to the HANG7 dataset is governed by data-use agreements, and it is therefore not publicly available.

## Code sharing

The models and source codes for reproducing the results reported in this paper can be accessed at https://github.com/yuty2009/sleepgpt.

## Declaration of interests

W.W. reports equity from Alto Neuroscience. None of the other authors has financial disclosures to report.

## Acknowledgements

This work was supported in part by STI2030-Major Projects under Grant 2022ZD0211700, AI Program of Shanghai Municipal Education Commission under Grant JWAIZD-4, the National Natural Science Foundation of China under Grant 62376098, 62276102, and U22A20293, and GuangDong Basic and Applied Basic Research Foundation 2024A1515011983.

## APPENDIX A Supplementary data

Additional study and data information is contained in the supplementary material.

## Notes

### Author Declarations

The study on the private HANG7 dataset was conducted at Zhejiang University with Institutional Review Board approval, and written consent was obtained from all participants or their caregivers.

### Summary of Updates

A revision for eLife submission.

## References

Association AP. Diagnostic and Statistical Manual of Mental Disorders, Fifth Edition, Text Revision (DSM-5-TR). Washington, DC, USA: American Psychiatric Publishing; 2022.

Born J, Manica M. Regression Transformer enables concurrent sequence regression and generation for molecular language modelling, Journal Article. Nature Machine Intelligence. 2023; 5(4):432–444. https://doi.org/10.1038/s42256-023-00639-z, doi: 10.1038/s42256-023-00639-z.

Dong H, Supratak A, Pan W, Wu C, Matthews PM, Guo Y. Mixed Neural Network Approach for Temporal Sleep Stage Classification, Journal Article. IEEE Transactions on Neural Systems and Rehabilitation Engineering. 2018; 26(2):324–333. https://www.ncbi.nlm.nih.gov/pubmed/28767373, doi: 10.1109/TNSRE.2017.2733220.

Eldele E, Chen Z, Liu C, Wu M, Kwoh CK, Li X, Guan C. An Attention-Based Deep Learning Approach for Sleep Stage Classification With Single-Channel EEG, Journal Article. IEEE Transactions on Neural Systems and Rehabilitation Engineering. 2021; 29:809–818. https://www.ncbi.nlm.nih.gov/pubmed/33909566, doi: 10.1109/TNSRE.2021.3076234.

Ghassemi MM, Moody BE, Lehman LH, Song C, Li Q, Sun H, Mark RG, Westover MB, Clifford GD. You Snooze, You Win: the PhysioNet/Computing in Cardiology Challenge 2018, Journal Article. Computing in Cardiology (2010). 2018; 45. doi: 10.22489/cinc.2018.049.

Goldberger AL, Amaral LAN, Glass L, Hausdorff JM, Ivanov PC, Mark RG, Mietus JE, Moody GB, Peng CK, Stanley HE. PhysioBank, PhysioToolkit, and PhysioNet. Circulation. 2000; 101(23):e215–e220. https://www. ahajournals.org/doi/abs/10.1161/01.CIR.101.23.e215, doi: 10.1161/01.CIR.101.23.e215.

Kemp B, Zwinderman AH, Tuk B, Kamphuisen HAC, Oberye JJL. Analysis of a sleep-dependent neuronal feedback loop: the slow-wave microcontinuity of the EEG. IEEE Transactions on Biomedical Engineering. 2000; 47(9):1185–1194. doi: 10.1109/10.867928.

Khalighi S, Sousa T, Santos JM, Nunes U. ISRUC-Sleep: A comprehensive public dataset for sleep researchers. Computer Methods and Programs in Biomedicine. 2016; 124:180–192. https://www.sciencedirect.com/science/article/pii/S0169260715002734, doi: 10.1016/j.cmpb.2015.10.013.

Lin Z, Akin H, Rao R, Hie B, Zhu Z, Lu W, Smetanin N, Verkuil R, Kabeli O, Shmueli Y, dos Santos Costa A, Fazel-Zarandi M, Sercu T, Candido S, Rives A. Evolutionary-scale prediction of atomic-level protein structure with a language model, Journal Article. Science. 2023; 379(6637):1123–1130. https://doi.org/10.1126/science.ade2574, doi: 10.1126/science.ade2574.

Lopez-Larraz E, Sierra-Torralba M, Clemente S, Fierro G, Oriol D, Minguez J, Montesano L, Klinzing JG. Bitbrain Open Access Sleep Dataset. . 2024; doi: 10.18112/openneuro.ds005555.v1.0.0.

Mahowald MW, Schenck CH. Insights from studying human sleep disorders, Journal Article. Nature. 2005; 437(7063):1279–85. https://www.ncbi.nlm.nih.gov/pubmed/16251953, doi: 10.1038/nature04287.

O’Reilly C, Gosselin N, Carrier J, Nielsen T. Montreal Archive of Sleep Studies: an open-access resource for instrument benchmarking and exploratory research. Journal of Sleep Research. 2014; 23(6):628–635. https://onlinelibrary.wiley.com/doi/abs/10.1111/jsr.12169, doi: 10.1111/jsr.12169.

Perslev M, Darkner S, Kempfner L, Nikolic M, Jennum PJ, Igel C. U-Sleep: resilient high-frequency sleep staging, Journal Article. npj Digital Medicine. 2021; 4(1):72. https://doi.org/10.1038/s41746-021-00440-5, doi: 10.1038/s41746-021-00440-5.

Phan H, Andreotti F, Cooray N, Chen OY, De Vos M. Joint Classification and Prediction CNN Framework for Automatic Sleep Stage Classification, Journal Article. IEEE Transactions on Biomedical Engineering. 2019; 66(5):1285–1296. https://www.ncbi.nlm.nih.gov/pubmed/30346277, doi: 10.1109/TBME.2018.2872652.

Phan H, Andreotti F, Cooray N, Chen OY, De Vos M. SeqSleepNet: End-to-End Hierarchical Recurrent Neural Network for Sequence-to-Sequence Automatic Sleep Staging, Journal Article. IEEE Transactions on Neural Systems and Rehabilitation Engineering. 2019; 27(3):400–410. https://www.ncbi.nlm.nih.gov/pubmed/30716040, doi: 10.1109/TNSRE.2019.2896659.

Phan H, Chen OY, Tran MC, Koch P, Mertins A, De Vos M. XSleepNet: Multi-View Sequential Model for Automatic Sleep Staging, Journal Article. IEEE Transactions on Pattern Analysis and Machine Intelligence. 2022; 44(9):5903–5915. https://www.ncbi.nlm.nih.gov/pubmed/33788679, doi: 10.1109/TPAMI.2021.3070057.

Phan H, Mikkelsen K, Chén OY, Koch P, Mertins A, De Vos M. SleepTransformer: Automatic Sleep Staging With Interpretability and Uncertainty Quantification. IEEE Transactions on Biomedical Engineering. 2022; 69(8):2456–2467. doi: 10.1109/TBME.2022.3147187.

Phyo J, Ko W, Jeon E, Suk HI. TransSleep: Transitioning-Aware Attention-Based Deep Neural Network for Sleep Staging. IEEE Transactions on Cybernetics. 2023; 53(7):4500–4510. doi: 10.1109/TCYB.2022.3198997.

Quan SF, Howard BV, Iber C, Kiley JP, Nieto FJ, O’Connor GT, Rapoport DM, Redline S, Robbins J, Samet JM, Wahl PW. The Sleep Heart Health Study: Design, Rationale, and Methods. Sleep. 1997; 20(12):1077–1085. https://doi.org/10.1093/sleep/20.12.1077, doi: 10.1093/sleep/20.12.1077.

Radford A, Narasimhan K, Salimans T, Sutskever I. Improving language understanding with unsupervised learning. . 2018; .

Radford A, Wu J, Child R, Luan D, Amodei D, Sutskever I, et al. Language models are unsupervised multitask learners. OpenAI blog. 2019; 1(8):9.

Rosenberg RS, Van Hout S. The American Academy of Sleep Medicine inter-scorer reliability program: sleep stage scoring, Journal Article. Journal of Clinical Sleep Medicine. 2013; 9(1):81–7. doi: 10.5664/jcsm.2350.

Seo H, Back S, Lee S, Park D, Kim T, Lee K. Intra- and inter-epoch temporal context network (IITNet) using sub-epoch features for automatic sleep scoring on raw single-channel EEG. Biomedical Signal Processing and Control. 2020; 61:102037. https://www.sciencedirect.com/science/article/pii/S1746809420301932, doi: 10.1016/j.bspc.2020.102037.

Stephansen JB, Olesen AN, Olsen M, Ambati A, Leary EB, Moore HE, Carrillo O, Lin L, Han F, Yan H, Sun YL, Dauvilliers Y, Scholz S, Barateau L, Hogl B, Stefani A, Hong SC, Kim TW, Pizza F, Plazzi G, et al. Neural network analysis of sleep stages enables efficient diagnosis of narcolepsy, Journal Article. Nature Communications. 2018; 9(1):5229. https://www.ncbi.nlm.nih.gov/pubmed/30523329, doi: 10.1038/s41467-018-07229-3.

Supratak A, Dong H, Wu C, Guo Y. DeepSleepNet: A Model for Automatic Sleep Stage Scoring Based on Raw Single-Channel EEG, Journal Article. IEEE Transactions on Neural Systems and Rehabilitation Engineering. 2017; 25(11):1998–2008. https://www.ncbi.nlm.nih.gov/pubmed/28678710, doi: 10.1109/TNSRE.2017.2721116.

Supratak A, Guo Y. TinySleepNet: An Efficient Deep Learning Model for Sleep Stage Scoring based on Raw Single-Channel EEG. In: The 42nd Annual International Conference of the IEEE Engineering in Medicine & Biology Society (EMBC); 2020. p. 641–644. doi: 10.1109/EMBC44109.2020.9176741.

Terzano MG, Parrino L, Sherieri A, Chervin R, Chokroverty S, Guilleminault C, Hirshkowitz M, Mahowald M, Moldofsky H, Rosa A, Thomas R, Walters A. Atlas, rules, and recording techniques for the scoring of cyclic alternating pattern (CAP) in human sleep. Sleep Medicine. 2001; 2(6):537–553. https://www.sciencedirect. com/science/article/pii/S1389945701001496, doi: 10.1016/S1389-9457(01)00149-6.

Thachayani M, Loganayagi M. Artificial intelligence based classifier for sleep disorder detec-tion using eeg-bci data. International Journal of Computer Science Trends and Technology. 2021; 9.

Vallat R, Walker MP. An open-source, high-performance tool for automated sleep staging, Journal Article. eLife. 2021; 10:e70092. doi: 10.7554/eLife.70092.

Wang J, Zhao S, Zhou Y, Jiang H, Yu Z, Li T, Li S, Pan G. Narcolepsy Diagnosis With Sleep Stage Features Using PSG Recordings. IEEE Transactions on Neural Systems and Rehabilitation Engineering. 2023; 31:3619–3629. doi: 10.1109/TNSRE.2023.3312396.

Xu S, Faust O, Seoni S, Chakraborty S, Barua PD, Loh HW, Elphick H, Molinari F, Acharya UR. A review of automated sleep disorder detection, Journal Article. Computers in Biology and Medicine. 2022; 150:106100. https://www.ncbi.nlm.nih.gov/pubmed/36182761, doi: 10.1016/j.compbiomed.2022.106100.

Zhang GQ, Cui L, Mueller R, Tao S, Kim M, Rueschman M, Mariani S, Mobley D, Redline S. The National Sleep Research Resource: towards a sleep data commons. Journal of the American Medical Informatics Association. 2018; 25(10):1351–1358. https://doi.org/10.1093/jamia/ocy064, doi: 10.1093/jamia/ocy064.

Zheng NS, Annis J, Master H, Han L, Gleichauf K, Ching JH, Nasser M, Coleman P, Desine S, Ruderfer DM, Hernandez J, Schneider LD, Brittain EL. Sleep patterns and risk of chronic disease as measured by long-term monitoring with commercial wearable devices in the All of Us Research Program, Journal Article. Nature Medicine. 2024; 30(9):2648–2656. https://doi.org/10.1038/s41591-024-03155-8, doi: 10.1038/s41591-024-03155-8.

