## Supplementary Material for "SleepGPT: A Sleep Stage Language Model for Efficient Sleep Assessment"

#### Datasets

This study utilizes three distinct data sets: sleep data sets for SleepGPT pretraining, sleep staging, and sleep disorder diagnosis. A total of eight publicly accessible sleep datasets are employed in the experiments. Table S1 provides a comprehensive summary of these datasets, and further details are provided below.

##### Datasets for SleepGPT pretraining

Training a transformer-based language model typically requires an extensive text corpus, often encompassing millions or even billions of web pages or documents. However, a sleep dataset of comparable scale, complete with sleep stage annotations, is not available. Fortunately, the increasing advancements in sleep medicine research, coupled with the research community's open data policy, have produced several publicly accessible sleep datasets with sleep stage annotations. The Sleep Heart Health Study (SHHS) database, a multi-center cohort study examining the cardiovascular and other consequences of sleep-disordered breathing ***Quan et al.*** (***1997***); ***Zhang et al.*** (***2018***), is a noteworthy example. This database comprises two rounds of PSG records: Visit 1 (SHHS-1) and Visit 2 (SHHS-2). In this work, we use the SHHS-1 cohort, which encompasses 5,793 subjects aged between 39 and 90 years, to train the SleepGPT model. Notably, to ensure alignment with the AASM scoring standard ***Iber et al.*** (***2007***), we merge the N3 and N4 stages into the N3 stage while discarding the MOVEMENT and UNKNOWN epochs, as the SHHS-1 database was manually scored following the R&K guidelines ***Wolpert*** (***1969***).

##### Datasets for sleep staging

The proposed SleepGPT model's performance on sleep staging tasks is evaluated with two widely used sleep datasets, namely, the Sleep-EDF and the Montreal Archive of Sleep Studies (MASS). The Physio2018 dataset serves as a benchmark to evaluate the generalization performance of SleepGPT in enhancing sleep staging. Table S2 provides a detailed summary of these datasets.

**SleepEDF Dataset**: We utilize the 2018 version of the SleepEDF Expanded dataset ***Kemp et al.*** (***2000***); ***Goldberger et al.*** (***2000***). This collection comprises data from 78 healthy Caucasian subjects aged 25 to 101 years. Each subject contributed two consecutive day-night PSG recordings, except for subjects 13, 36, and 52, where one recording was lost due to device failure. Consequently, the dataset contains 153 overnight recordings. Sleep experts manually scored the epochs based on the R&K standard ***Wolpert*** (***1969***), assigning each 30-second PSG epoch to one of eight categories: {W, N1, N2, N3, N4, REM, MOVEMENT, UNKNOWN}. To align with convention, the N3 and N4 stages were merged into the N3 stage, while the MOVEMENT and UNKNOWN epochs were excluded. Notably, the SleepEDF-20 dataset was not assessed because it is a subset of this particular version of SleepEDF.

**MASS Dataset**: Derived from different hospital-based sleep laboratories, the MASS database comprises whole-night recordings from 200 subjects (97 males and 103 females) aged 18 to 76 years ***O'Reilly et al.*** (***2014***). The annotation process involved sleep experts adhering to either the AASM standard ***Iber et al.*** (***2007***) (for the SS1 and SS3 subsets) or the R&K standard ***Wolpert*** (***1969***) (for the SS2, SS4, and SS5 subsets). In alignment with the previously mentioned datasets, we harmonized the R&K annotations with the five sleep stages {W, N1, N2, N3, REM} according to the AASM standard. Epochs initially spanning 20 seconds were extended to 30 seconds by incorporating the 5-second segments before and after them.

**Physio2018 Dataset**: The PhysioNet 2018 Challenge dataset, also known as the Physio2018 dataset, comprises 1,985 polysomnographic recordings provided by the Computational Clinical Neurophysiology Laboratory (CCNL) and the Clinical Data Animation Center (CDAC) at Massachusetts General Hospital (MGH). This dataset was used in the 2018 PhysioNet Challenge ***Ghassemi et al.*** (***2018***); ***Goldberger et al.*** (***2000***) to detect sleep arousals. We used the training set for our experiments, which included 944 subjects aged 18 to 90. Sleep experts manually scored the recordings according to the American Academy of Sleep Medicine (AASM) guidelines ***Iber et al.*** (***2007***), annotating five sleep stages: W, N1, N2, N3, and REM. This dataset is employed to blindly validate the sleep staging model derived from the MASS dataset.

**BOAS Dataset**: The Bitbrain Open Access Sleep (BOAS) dataset serves to bridge the gap between gold-standard clinical sleep monitoring and emerging wearable EEG technologies ***Lopez-Larraz et al.*** (***2024***). This dataset comprises data from 128 nights, during which healthy participants were simultaneously monitored using both a Brain Quick Plus Evolution PSG system by Micromed and a Bitbrain wearable EEG headband. The Micromed PSG system collected EEG signals from electrodes placed at F3, F4, C3, C4, O1, and O2, following the international 10-20 system. In contrast, the Bitbrain headband recorded EEG signals from the frontal AF7 and AF8 electrode sites. Both systems utilized a sampling rate of 256 Hz. Sleep staging was independently annotated by three expert scorers following the American Academy of Sleep Medicine (AASM) criteria ***Iber et al.*** (***2007***), with a consensus label established by a fourth expert. The BOAS dataset enables the evaluation of the SleepGPT-based model's ability to achieve sleep staging accuracy comparable to PSG using wearable EEG data.

#### Datasets for sleep disorder diagnosis

Two sleep-disorder-related datasets were identified for the sleep disorder diagnosis analysis, including the CAP Sleep Database and the Mignot Nature Communications (MNC) dataset, which offer both sleep stage annotations and sleep disorder labels.

**CAP Dataset**: The CAP Sleep Database is a collection of 108 polysomnographic recordings contributed by the Sleep Disorders Center of the Ospedale Maggiore of Parma, Italy ***Terzano et al.*** (***2001***); ***Goldberger et al.*** (***2000***). It contains data from seven groups of patients with distinct sleep disorders, as well as a healthy control group without any medical, neurological, or psychiatric conditions. The details of these groups are summarized in Table S3. Well-trained neurologists who are sleep experts manually scored the sleep recordings based on the R&K rules, categorizing the epochs into sleep stages 1-4, wake, REM sleep, and movement artifacts. To adhere to the AASM standard, we harmonized the R&K annotations, obtaining five sleep stages: {W, N1, N2, N3, REM}. Notably, the CAP Sleep Database has been widely used in numerous studies focusing on sleep disorder diagnosis.

Note that the CAP Sleep Database is inherently imbalanced. Several groups contain an extremely limited number of subjects. For instance, the BRUX and SBD groups consisted of only 2 and 3 subjects, respectively. To facilitate subject-level diagnoses, we conducted binary classification experiments on the CAP Sleep Database, specifically distinguishing abnormal from normal cases. Furthermore, after incorporating the first-session data of 78 subjects (excluding subjects 36 and 52, who had only one session available) from the SleepEDF Expanded database into the normal group, the comprehensive dataset included 186 subjects, 94 of whom were labeled as normal and 92 of whom were labeled as abnormal.

**ISRUC Dataset**: The ISRUC-Sleep dataset ***Khalighi et al.*** (***2016***) contains full-night PSG recordings, each approximately eight hours in duration, collected at the Sleep Medicine Centre of Coimbra University Hospital (CHUC) between 2009 and 2013. Data were acquired non-invasively using a SomnoStar Pro multi-channel system, with sensors placed according to the international 10-20 standard. The dataset includes recordings from both healthy adults and subjects with sleep disorders under medication, divided into three groups: 1) 100 subjects with one session each; 2) 8 subjects with two sessions for longitudinal studies; 3) 10 healthy subjects with one session, used for comparison with sleep disorder patients. Among the dataset, 76 obstructive sleep apnea (OSA) patients and 10 healthy controls are included to blindly validate the sleep disorder diagnosis model derived from the CAP dataset.

**MNC Dataset**: The Mignot Nature Communications (MNC) dataset comprises raw polysomnography data collected from an automated sleep staging project utilizing neural networks ***Zhang et al.*** (***2018***); ***Stephansen et al.*** (***2018***). It encompasses data from ten distinct cohorts recorded at twelve sleep centers across three continents: the patient-based Stanford Sleep Cohort (SSC), the population-based Wisconsin Sleep Cohort (WSC), the patient-based Inter-scorer Reliability Cohort (IS-RC), the Jazz Clinical Trial Sample (JCTS), the patient-based Korean Hypersomnia Cohort (KHC), the patient-based Austrian Hypersomnia Cohort (AHC), the patient-based Italian Hypersomnia Cohort (IHC), the patient-based Danish Hypersomnia Cohort (DHC), the patient-based French Hypersomnia Cohort (FHC), and the patient-based Chinese Narcolepsy Cohort (CNC). The study received approval from institutional review boards, and informed consent was obtained from all participants. Trained sleep-scoring technicians manually annotated all sleep studies according to the AASM Scoring Manual ***Iber et al.*** (***2007***). Additionally, the subjects were divided into the Type-1 narcolepsy (T1N; with either low CSF hypocretin-1 levels or clear cataplexy), other hypersomnia (OHS), or non-narcolepsy control (NNC) group based on multiple sleep latency test (MSLT) results, cataplexy symptoms, and human leukocyte antigen (HLA) results (if available). Further information about the MNC dataset can be found in ***Stephansen et al.*** (***2018***).

However, as diagnostic results were only available for the SSC, DHC, and CNC cohorts, we exclusively utilized these three cohorts for the sleep disorder diagnosis task. The dataset compiled from these cohorts encompassed 407 subjects, among whom 82 were in the T1N group, 43 were in the OHS group, and 282 were in the NNC group (see Table S4). To facilitate comparison with previous research, we adhered to the methodology outlined in ***Stephansen et al.*** (***2018***) and conducted binary classification experiments on the MNC dataset to discriminate T1N subjects from all other subjects, i.e., OHS and NNC subjects. This dataset is also employed to blindly assess the generalizability of the sleep disorder diagnosis model derived from the CAP dataset.

**HANG7 Dataset**: In the HANG7 dataset ***Wang et al.*** (***2023***), 84 participants aged 11 to 57 years (mean age: 24.5 $\pm$ 9.6), including 44 females and 40 males, were recruited to collect polysomnography recordings at the Affiliated Mental Health Center & Hangzhou Seventh People's Hospital, Zhejiang University School of Medicine. This study was conducted in accordance with the Declaration of Helsinki and approved by the Institutional Review Board of the Ethics Committee of Hangzhou Seventh People's Hospital, China (Approval No. 2023-053-02). Written informed consent was obtained from all participants or their legal caregivers prior to participation. PSG recordings were collected following the AASM sleep standards ***Iber et al.*** (***2007***) and manually scored by experienced sleep technicians. Each participant's PSG recording covered one full night, from approximately 21:00 to 5:00 the next morning, totaling about 8 hours. Of the 84 participants, 13 were diagnosed with T1N (with clear cataplexy), 38 were other narcolepsy, and 33 were healthy controls. The dataset was used to blindly validate the generalizability of the sleep disorder diagnosis models derived from the CAP (abnormal vs. normal) and MNC (T1N vs. others) datasets.

#### Methods in detail

##### Problem formulation

###### Sleep staging as speech recognition

A general speech recognition system consists of an acoustic model and a language model. The acoustic model $\mathcal{M}_1$ predicts the most likely word sequence given speech audio features $X$ via

$$P(Y; \mathcal{M}_1) = P(Y|X; \mathcal{M}_1), \tag{S1}$$

while the language model $\mathcal{M}_2$ is built based on a large-scale text corpus to capture the sequential characteristics of the word sequence via

$$P(Y; \mathcal{M}_2) = P(y_1, y_2, \cdots, y_T) = \prod_{t=1}^{T} P(y_t|y_1, y_2, \cdots, y_{t-1}). \tag{S2}$$

The trained language model is then used to rectify the word sequence predicted by the acoustic model to improve the speech recognition performance as follows ***Chan et al.*** (***2016***); ***Rao et al.***

(***2017***); ***Karita et al.*** (***2019***):

$$\begin{aligned} Y^{\star} &= \arg\max_{Y} P(Y; \mathcal{M}_1, \mathcal{M}_2) \\ &= \arg\max_{Y} P(Y|X; \mathcal{M}_1) P(Y; \mathcal{M}_2). \end{aligned} \quad \text{(S3)}$$

In the automated sleep staging process, features are extracted from a sleep data epoch, and a sleep staging model is then applied to predict the most likely sleep stage for that epoch. The model, whether it is a traditional machine learning model or a deep learning model, is similar to the acoustic model in a speech recognition system. It is designed and trained on a large-scale sleep dataset to achieve satisfactory prediction accuracy on unseen sleep epochs. However, due to various factors such as architectural limitations, training inadequacies, and data scarcity, imperfections in the PSG-based sleep staging model may occur, leading to inaccurate predictions of sleep stages. The inter-scorer reliability for sleep stage scoring is reported to be 82.6% on average, which closely aligns with that achieved by machine learning-based automated staging systems ***Stephansen et al.*** (***2018***). Given the analogous nature of text and sleep stage sequences, emulating the speech recognition system paradigm may improve sleep staging performance. This involves training a sleep language model that can capture the inherent sequential characteristics of sleep stages to assume a role akin to that of a natural language model (see Fig. **2B**).

##### Sleep disorder diagnosis as text classification

It is believed that sleep architecture, i.e., the distribution of sleep stages, is strongly related to the quality of sleep. Certain sleep disorders exhibit disrupted sleep architecture and atypical sleep stage transitions. Examples include shortened deep sleep stages in cases of insomnia, and short REM sleep latency or immediate transitions from wakefulness to REM sleep in narcolepsy. An overnight sleep stage sequence has the form $\{\mathrm{W}, \cdots, \mathrm{N1}, \cdots, \mathrm{N2}, \cdots, \mathrm{N3}, \cdots, \mathrm{REM}, \cdots\}$. Such sequences are plotted as hypnograms in Fig. **1** (top-left). They are akin to concise news pieces or textual documents with a vocabulary of only five words. In turn, the sleep stage sequence encodes the sequential attributes and transitional patterns characterizing the progression between sleep stages. The integration of a sequential model enables these characteristics to be identified and described.

Hence, a viable approach is to treat sleep disorder diagnosis as a long text classification task. Here, the trained sleep language model is harnessed to extract features from the sleep stage sequences (hypnograms), enabling the subsequent classification of these sequences as associated with a sleep disorder or not. This is an innovative method for sleep disorder diagnosis using only the sleep stages obtained from either human experts or automated sleep staging models.

#### The SleepGPT model

Recently, the success of transformer-based language models (LMs), including bidirectional encoder representations from transformers (BERT) and generative pretrained transformers (GPT), has been noted across various natural language processing (NLP) tasks, such as machine translation, question-answering, and text generation ***Kenton and Toutanova*** (***2019***); ***Radford et al.*** (***2018***, 2019). BERT, which functions as a bidirectional LM, is trained to predict masked words based on neighboring words and to perform next-sentence prediction. Conversely, GPT operates as a decoder-only transformer LM, autoregressively predicting the next character or word based on preceding ones. Both models undergo self-supervised training on extensive text corpora. The success of ChatGPT has illustrated its proficient ability to capture the inherent sequential attributes of natural language. This ability has been extensively utilized to improve performance in speech recognition and diverse NLP tasks, such as text classification.

Transformers consist of multiple transformer blocks, typically including a multi-head self-attention layer, a feed-forward layer, and residual and normalization layers ***Vaswani et al.*** (***2017***). GPT employs causal attention in its self-attention layer to ensure exclusive attention to preceding words.

This concept has been applied in sequential sleep staging models that rely on prior sleep epochs for current sleep stage predictions. It has proven to be especially significant in online sleep staging systems, where only preceding epochs are accessible during the current prediction task. Hence, GPT is selected to capture the sequential traits of sleep stages. The SleepGPT model adopts the architecture of the GPT-2 language model developed by OpenAI ***Radford et al.*** (***2018***, 2019). As illustrated in Fig. **2A**, the main part of the model is composed of a series of $n$ transformer decoder blocks, marked by the masked multi-head self-attention layers. Token embedding and position embedding layers are employed to map the sleep stage tokens into a vector space and to infuse positional information into each token's sequence placement, respectively. The final transformer block's output is input to a linear layer to obtain the probability distribution of the next sleep stage.

The model undergoes self-supervised training through the autoregressive forecast of the most likely sleep stage based on preceding sleep stages. For each subject or sleep session, the overnight sleep stage annotations are structured as a long sequence $\mathcal{U} = \{u_1, ..., u_N\}$, where $u_i \in \{0, 1, 2, 3, 4\}$ represents the five sleep stages $\{\mathrm{W}, \mathrm{N1}, \mathrm{N2}, \mathrm{N3}, \mathrm{REM}\}$ as integers ranging from 0 to 4. Training samples are derived from the sequence using a sliding window of dimensions $K$, passing over the sequence with a stride of 1, where $K$ signifies the sample sequence length. Consequently, the overnight stage sequence yields $N - K + 1$ overlapping instances. The $(i-1)$th instance of the $K$ sleep stages, denoted as $\mathbf{U}_{i-1} = \{u_{i-K}, ..., u_{i-1}\}$, constitutes the input and is fed into the model for predicting the target; the target is the $i$th instance $\mathbf{U}_i = \{u_{i-K+1}, ..., u_i\}$:

$$h_0 = \mathbf{U}_{i-1}\mathbf{W}_e + \mathbf{W}_p \quad \text{(S4)}$$

$$h_l = \text{transformer\_block}(h_{l-1}),\ \forall l \in [1, L] \quad \text{(S5)}$$

$$P(\mathbf{U}_i) = \text{softmax}(h_n \mathbf{W}_s^T) \quad \text{(S6)}$$

where $L$ is the number of transformer layers, $\mathbf{W}_e$ is the token embedding matrix, $\mathbf{W}_p$ is the position embedding matrix, and $\mathbf{W}_s$ is the weight of the classification head. The model is trained with the objective of minimizing the cross-entropy loss, thereby reducing the disparity between the predicted sleep stage and the actual ground truth:

$$\mathcal{L}(\mathcal{U}) = -\sum_{i=1} \log P(\mathbf{U}_i | \mathbf{U}_{i-1}; \Theta), \quad \text{(S7)}$$

where $\Theta$ is the parameter of the GPT sequential model.

#### Sleep staging with SleepGPT

Automated sleep staging is an active research topic within the realm of sleep medicine; it aims to automate the prediction of sleep stages for individual sleep PSG data epochs, typically spanning 30 seconds. Several excellent deep learning models have been proposed for this purpose. Most of them share a common architecture, comprising a CNN to extract intra-epoch features and an RNN to incorporate the contextual information in adjacent PSG data epochs***Supratak et al.*** (***2017***); ***Phan et al.*** (***2019a***); ***Mousavi et al.*** (***2019***); ***Phan et al.*** (***2019b***); ***Supratak and Guo*** (***2020***); ***Phan et al.*** (***2022***). Among cutting-edge sleep staging models, XSleepNet***Phan et al.*** (***2022***) employs two network streams to learn from multi-view inputs (e.g., both raw signals and time-frequency images) for sleep staging. By adapting the contributions of the two views on time to perform joint feature learning during training, XSleepNet outperforms the single-view baselines and multi-view baselines with a simple fusion strategy. However, while contextual information within PSG signals is incorporated into the above deep learning models, they neglect the inherent sequential traits and transition patterns within sleep stages.

To use the trained SleepGPT model to improve sleep staging performance, we treat the sleep staging task as a speech recognition task and follow the pipeline in Fig. **2B**. The sleep staging model (SSM) is used to predict the most likely sleep stage given a PSG data epoch or preceding epochs, while the SleepGPT model is used to rectify the predicted sleep stage given past stages. That is, the output logits of the SSM $P_{\text{SSM}}(y)$ and those of the sleep language model (SLM) $P_{\text{SLM}}(y)$ are weighted by a factor $\alpha$ to obtain the final sleep stage prediction:

$$P(y) = \alpha P_{\text{SSM}}(y|\mathbf{x}) + (1-\alpha)P_{\text{SLM}}(y|\mathbf{y}_{-}), \tag{S8}$$

where $\mathbf{x}$ is the set of input data epochs, including the current and preceding epochs depending on whether a memory staging model is used, and $\mathbf{y}_{-}$ is the set of preceding sleep stages (context). Notably, the hyperparameter $\alpha$ governs the relative influences of these two models, ultimately steering the sleep stage prediction toward the highest probability outcome.

#### Sleep disorder diagnosis with SleepGPT

Sleep disorder diagnosis involves discerning the presence and specific type of sleep disorder in an individual. This task is typically conducted either by sleep specialists or by ML models that leverage PSG data for automated assessment. ML-based approaches often include PSG data feature extraction, followed by the classification of sleep disorders. However, the high dimensionality of PSG data, coupled with a lack of labeled instances, makes it challenging to train an ML model with robust generalizability. The situation is even worse in the case of deep learning models, which commonly demand an extensive volume of labeled data to achieve optimal model performance.

Pretrained language models, such as GPTs trained on expansive text corpora, can capture the intrinsic sequential characteristics of natural language. These pretrained sleep language models can function as feature extractors, processing text sequences to subsequently enable classification. Notably, these pretrained models can be further fine-tuned using limited-scale datasets, significantly enhancing the classification performance. This strategy, referred to as transfer learning, effectively addresses the challenge of limited sample size, and it has a proven track record of success across numerous NLP applications.

Drawing inspiration from this pretraining and fine-tuning paradigm, we leverage the pretrained SleepGPT model as a feature extractor. We replace the subsequent stage prediction layer with a classifier and perform comprehensive fine-tuning to facilitate sleep disorder classification. Nevertheless, a challenge arises for the SleepGPT model in handling long sequences: the sequence length limitation. A whole-night (8-hour) sleep stage sequence comprises 960 time steps, significantly exceeding the sequence length restriction of the SleepGPT model. We address this by segmenting the overnight sequence into shorter sections. Initially, the SleepGPT model is employed to extract local context features from these short sleep stage segments. Subsequently, another sequential model (a transformer encoder) is used to capture the global contextual features from the SleepGPT output ***Vaswani et al.*** (***2017***). Finally, the resulting global context features are fed into a classification head to predict sleep disorder labels. The configuration of the hierarchical transformer network (HTN) is depicted in Fig. S1. This architecture, which is tailored for lengthy sequence classification, is borrowed from the hierarchical attention network (HAN) and hierarchical transformers utilized for long text classification ***Yang et al.*** (***2016***); ***Pappagari et al.*** (***2019***).

Additionally, to facilitate mini-batch training, we pad the short sleep stage sequences within a batch to achieve a uniform length. Subsequently, a mask matrix is employed to exclude the padded values during the computation of attention weights and the loss calculation. The loss function involves cross-entropy loss, which compares the predicted sleep disorder labels with the ground truth:

$$\mathcal{U} = \{\mathbf{U}_i\},\ \forall i \in [1, N] \tag{S9}$$

$$\mathbf{z}_i = \text{SleepGPT}(\mathbf{U}_i) \tag{S10}$$

$$\mathbf{V} = \text{transformer\_encoder}(\mathbf{Z}) \tag{S11}$$

$$P(\mathcal{U}) = \text{softmax}(\mathbf{W}_c \mathbf{v}_0 + b_c) \tag{S12}$$

$$\mathcal{L}(\mathcal{U}) = -\sum_{i=1} \log P(\mathcal{U}; \Phi), \tag{S13}$$

where $\mathbf{U}_i$ is the $i$th segment, and $N$ represents the number of segments within a sleep stage sequence. $\mathbf{z}_i$ denotes the $i$th local feature vector generated by SleepGPT. Notably, $\mathbf{Z} = \{\mathbf{z}_0, \cdots, \mathbf{z}_N\}$ and $\mathbf{V} = \{\mathbf{v}_0, \cdots, \mathbf{v}_N\}$ denote the input and output sequences of the transformer encoder, respectively. Here, $\mathbf{z}_0$ corresponds to the $CLS$ token, while $\mathbf{v}_0$ represents the global feature vector at the $CLS$ token's output position. The $CLS$ token in transformers is a special token added at the beginning of the input sequence, and its final output serves as a global representation of the entire sequence for tasks like classification ***Kenton and Toutanova*** (***2019***); ***Dosovitskiy et al.*** (***2021***). The parameters of the classification head are denoted as $\mathbf{W}_c$ and $b_c$, and $\Phi$ includes the trainable parameters of the entire model.

#### Implementation details

All models are implemented in PyTorch using the HuggingFace Transformers framework. Training and evaluation are performed on NVIDIA GPUs. Unless otherwise specified, optimization is carried out using the Adam optimizer with standard momentum parameters.

Hyperparameter settings are selected based on validation performance within the training data only. For SleepGPT pretraining, a fixed set of hyperparameters is used across all experiments, including the learning rate, batch size, and training epochs. For downstream sleep staging and sleep disorder diagnosis tasks, the same hyperparameter configurations are applied across datasets whenever possible to avoid dataset-specific tuning. When minor adjustments are required due to differences in dataset size or task formulation, these adjustments are determined exclusively on the corresponding training or validation splits.

Where applicable, data splitting is performed at the subject level, ensuring no overlap between training, validation, and testing sets.

### Supplementary tables

**Table S1.** Overview of the involved sleep datasets. **BMI** = body mass index, **AHI** = apnea-hypopnea index.

| Dataset | Subjects / Sessions | Recording Duration | Age | Sex (% Male) | BMI | AHI | Health Conditions |
|---|---|---|---|---|---|---|---|
| **SHHS** *Quan et al.* (*1997*); *Zhang et al.* (*2018*) | 5793 / 5793 | Overnight | 63.1 ± 11.2 | 47.6 | 28.2 ± 5.1 | 17.9 ± 16.1 | Sleep-disordered breathing, heart diseases, and others |
| **SleepEDF** *Kemp et al.* (*2000*); *Goldberger et al.* (*2000*) | 78 / 153 | Around 9h | 59 ± 22.1 | 46.4 | - | - | Healthy subjects |
| **MASS** *O'Reilly et al.* (*2014*) | 200 / 200 | Overnight | 40.6 ± 19.4 | 48.5 | - | ≤ 20 | Healthy subjects |
| **Physio2018** *Ghassemi et al.* (*2018*); *Goldberger et al.* (*2000*) | 994 / 994 | 7.7h | 55 ± 14.3 | 67.0 | 33 ± 7.8 | 19 ± 14.6 | Sleep disorders, healthy subjects |
| **BOAS** *Lopez-Larraz et al.* (*2024*) | 128 / 128 | Overnight | 42.2 ± 19.0 | 40.6 | 23.8 ± 3.2 | - | Healthy subjects |
| **CAP** *Terzano et al.* (*2001*); *Goldberger et al.* (*2000*) | 108 / 108 | 8–10h | 45.2 ± 19.7 | 61.1 | - | - | Sleep disorders (N=92), healthy subjects (N=16) |
| **ISRUC** *Khalighi et al.* (*2016*) | 86 / 86 | Overnight | 49.7 ± 15.7 | 59.4 | - | - | Sleep apnea (N=76), healthy subjects (N=10) |
| **MNC-CNC** *Zhang et al.* (*2018*); *Stephansen et al.* (*2018*) | 77 / 77 | Overnight | 28.5 ± 16.9 | 51.3 | 23.2 ± 11.5 | 5.34 ± 1.51 | Type-1 narcolepsy (N=54), healthy subjects (N=23) |
| **MNC-DHC** *Zhang et al.* (*2018*); *Stephansen et al.* (*2018*) | 79 / 79 | Overnight | 33.4 ± 14.8 | 50.0 | 24.8 ± 4.9 | - | Type-1 narcolepsy (N=21), hypersomnia (N=38), healthy subjects (N=20) |
| **MNC-SSC** *Zhang et al.* (*2018*); *Stephansen et al.* (*2018*) | 251 / 251 | Overnight | 45.4 ± 13.8 | 59.4 | 23.9 ± 6.5 | 13.7 ± 0.7 | Type-1 narcolepsy (N=7), hypersomnia (N=5), healthy subjects (N=239) |
| **HANG7** *Wang et al.* (*2023*) | 84 / 84 | 8h | 24.5 ± 9.6 | 47.6 | 22.72 ± 3.65 | - | Type-1 narcolepsy (N=13), other narcolepsy (N=38), healthy subjects (N=33) |

**Table S2.** Number of subjects, EEG channels, and sleep stage distribution of the sleep staging datasets.

| Datasets | Subjects | EEG channel | W | N1 | N2 | N3 | REM | Total |
|---|---|---|---|---|---|---|---|---|
| SleepEDF | 78 | Fpz-Cz | 69824 | 21522 | 69132 | 13039 | 25835 | 199352 |
| MASS | 200 | C4-A1/C3-A2 | 31184 | 19359 | 107930 | 30383 | 40184 | 229040 |
| Physio2018 | 994 | C3-A2 | 157945 | 136978 | 377870 | 102592 | 116877 | 892262 |
| BOAS | 128 | C4/AF7 | 19137 | 4462 | 72181 | 5225 | 18754 | 120095 |

**Table S3.** Sleep disorder groups in the CAP Sleep Database.

| Sleep disorder | No. of subjects |
|---|---|
| Bruxism (BRUX) | 2 |
| Sleep-disordered breathing (SBD) | 4 |
| Insomnia (INS) | 9 |
| Narcolepsy (NARCO) | 5 |
| Nocturnal frontal lobe epilepsy (NFLE) | 40 |
| Periodic leg movements (PLMs) | 10 |
| REM behavior disorder (RBD) | 22 |
| No pathology (N) | 16 |

**Table S4.** Sleep disorder groups in the MNC dataset.

| Sleep disorder | No. of subjects | | | |
|---|---|---|---|---|
| | CNC | DHC | SSC | Total |
| Type-1 narcolepsy (T1N) | 54 | 21 | 7 | 82 |
| Other hypersomnia (OHS) | 0 | 38 | 5 | 43 |
| Non-narcolepsy control (NNC) | 23 | 20 | 239 | 282 |
| Total | 77 | 79 | 251 | 407 |

**Table S5.** Results of three state-of-the-art staging methods with and without SleepGPT when performing cross-dataset sleep staging on the SleepEDF ***Kemp et al.*** (***2000***), MASS ***O'Reilly et al.*** (***2014***), and Physio2018 ***Ghassemi et al.*** (***2018***) datasets. **Dataset**: **Source** → **Target** indicate that the staging model is trained from the **Source** dataset and evaluated on the **Target** dataset.

| Method | Dataset | SleepGPT | ACC | MF1 | kappa | W | N1 | N2 | N3 | REM |
|---|---|---|---|---|---|---|---|---|---|---|
| YASA | YASA → SleepEDF | w/o | 72.7 | 63.3 | 0.611 | 92.6 | 13.2 | 73.9 | 73.2 | 64.9 |
| | | w | 76.9 | 68.2 | 0.677 | 89.1 | 20.2 | 81.3 | 81.4 | 76.8 |
| | YASA → MASS | w/o | 78.6 | 71.2 | 0.700 | 92.8 | 21.7 | 82.0 | 91.6 | 84.3 |
| | | w | 80.2 | 73.6 | 0.724 | 95.4 | 26.1 | 82.3 | 94.0 | 86.5 |
| | YASA → Physio2018 | w/o | 71.4 | 66.2 | 0.613 | 92.5 | 20.5 | 77.3 | 78.1 | 77.4 |
| | | w | 73.5 | 68.6 | 0.640 | 93.6 | 22.8 | 80.7 | 79.3 | 77.7 |
| TinySleepNet | Physio2018 → MASS | w/o | 74.1 | 67.1 | 0.617 | 66.1 | 27.4 | 89.2 | 74.1 | 69.6 |
| | | w | 77.0 | 71.3 | 0.665 | 66.2 | 39.1 | 90.1 | 82.9 | 71.8 |
| | MASS → Physio2018 | w/o | 70.5 | 66.3 | 0.600 | 94.7 | 35.0 | 75.9 | 48.6 | 81.6 |
| | | w | 72.1 | 68.0 | 0.621 | 95.1 | 36.4 | 78.2 | 51.3 | 81.8 |
| XSleepNet | Physio2018 → MASS | w/o | 74.4 | 68.5 | 0.626 | 64.3 | 39.1 | 88.6 | 83.0 | 61.4 |
| | | w | 76.7 | 71.3 | 0.661 | 68.8 | 43.0 | 90.0 | 86.5 | 62.0 |
| | MASS → Physio2018 | w/o | 70.3 | 65.8 | 0.594 | 94.2 | 41.8 | 77.1 | 38.8 | 77.0 |
| | | w | 72.2 | 67.4 | 0.617 | 95.0 | 42.9 | 80.6 | 39.0 | 77.8 |

**Table S6.** Performance of state-of-the-art sleep staging methods and the proposed SleepGPT-powered models on the BOAS wearable dataset ***Lopez-Larraz et al.*** (***2024***). The best results are highlighted in bold, while the results with the SleepGPT-powered model are gray-shaded.

| Dataset | Method | SleepGPT | ACC | MF1 | kappa | W | N1 | N2 | N3 | REM |
|---|---|---|---|---|---|---|---|---|---|---|
| PSG | TinySleepNet | w/o | 80.9 | 64.1 | 0.660 | 78.0 | 20.8 | 90.8 | 48.0 | 69.4 |
| | | w | 81.6 | 65.6 | 0.673 | 78.4 | 23.1 | **91.2** | 51.1 | 70.5 |
| | XSleepNet | w/o | 83.4 | 68.8 | 0.715 | 88.7 | 27.0 | 88.2 | 53.9 | 81.1 |
| | | w | **84.3** | **70.4** | **0.728** | **89.0** | **27.9** | 89.2 | **56.3** | **81.7** |
| Headband | TinySleepNet | w/o | 80.7 | 62.5 | 0.652 | 78.5 | 18.3 | 91.1 | 39.5 | 68.8 |
| | | w | 81.3 | 64.5 | 0.667 | 79.5 | 20.8 | 91.0 | **48.0** | 69.4 |
| | XSleepNet | w/o | 83.0 | 65.7 | 0.694 | 84.7 | 23.9 | 91.8 | 36.8 | 73.4 |
| | | w | **83.9** | **67.8** | **0.712** | **87.0** | **26.9** | **91.9** | 43.8 | **74.5** |

**Table S7.** Performance of SleepGPT on sleep disorder diagnosis across datasets. The model was evaluated using cross-validation on the CAP dataset ***Terzano et al.*** (***2001***) and external validation on the ISRUC ***Khalighi et al.*** (***2016***), MNC ***Mignot et al.*** (***2002***), and HANG7 ***Wang et al.*** (***2023***) datasets for abnormal versus normal sleep classification.

| Dataset | Method | BACC | SENS | SPEC |
|---|---|---|---|---|
| **CAP (CV)** | Hypnogram | $0.910_{(0.868–0.948)}$ | $0.957_{(0.912–0.990)}$ | $0.863_{(0.787–0.931)}$ |
| | BaseNet | $0.849_{(0.793–0.899)}$ | $0.868_{(0.796–0.930)}$ | $0.830_{(0.750–0.905)}$ |
| | From scratch | $0.893_{(0.848–0.935)}$ | $0.902_{(0.835–0.957)}$ | $0.884_{(0.816–0.947)}$ |
| | Pretrained | $\mathbf{0.962}_{(0.935–0.985)}$ | $\mathbf{0.989}_{(0.962–1.000)}$ | $\mathbf{0.936}_{(0.884–0.979)}$ |
| **CAP → ISRUC** | Hypnogram | $0.804_{(0.638–0.945)}$ | $0.908_{(0.846–0.962)}$ | $0.700_{(0.375–1.000)}$ |
| | BaseNet | $0.670_{(0.504–0.830)}$ | $0.842_{(0.759–0.921)}$ | $0.498_{(0.182–0.800)}$ |
| | From scratch | $0.766_{(0.599–0.921)}$ | $0.934_{(0.877–0.987)}$ | $0.597_{(0.250–0.909)}$ |
| | Pretrained | $\mathbf{0.899}_{(0.750–1.000)}$ | $\mathbf{1.000}_{(1.000–1.000)}$ | $\mathbf{0.797}_{(0.500–1.000)}$ |
| **CAP → MNC** | Hypnogram | $0.778_{(0.735–0.820)}$ | $0.825_{(0.756–0.889)}$ | $0.731_{(0.675–0.781)}$ |
| | BaseNet | $0.749_{(0.697–0.806)}$ | $0.671_{(0.570–0.778)}$ | $0.828_{(0.786–0.864)}$ |
| | From scratch | $0.800_{(0.758–0.842)}$ | $0.841_{(0.771–0.902)}$ | $0.758_{(0.708–0.805)}$ |
| | Pretrained | $\mathbf{0.837}_{(0.801–0.875)}$ | $\mathbf{0.879}_{(0.825–0.929)}$ | $\mathbf{0.795}_{(0.749–0.841)}$ |
| **CAP → HANG7** | Hypnogram | $0.796_{(0.703–0.883)}$ | $0.803_{(0.686–0.907)}$ | $0.789_{(0.640–0.921)}$ |
| | BaseNet | $0.730_{(0.629–0.824)}$ | $0.823_{(0.709–0.922)}$ | $0.637_{(0.469–0.793)}$ |
| | From scratch | $0.769_{(0.671–0.859)}$ | $0.748_{(0.630–0.860)}$ | $0.791_{(0.628–0.923)}$ |
| | Pretrained | $\mathbf{0.840}_{(0.750–0.917)}$ | $\mathbf{0.864}_{(0.765–0.946)}$ | $\mathbf{0.815}_{(0.667–0.939)}$ |

Values are reported as point estimates with 95% confidence intervals (shown as subscripts). Confidence intervals were estimated using bootstrap resampling. **BACC**, balanced accuracy; **SENS**, sensitivity; **SPEC**, specificity. **Hypnogram** and **Hypnodensity** denote empirical feature-based XGBoost classifiers. **BaseNet** denotes a baseline neural network. **From scratch** refers to the hierarchical transformer network (HTN) trained without pretraining. **Pretrained** denotes the HTN initialized with pretrained SleepGPT parameters. Bold values indicate the best performance for each dataset.

**Table S8.** Performance of SleepGPT on narcolepsy diagnosis across datasets. The model was evaluated using cross-validation on the MNC dataset ***Mignot et al.*** (***2002***) and external validation on the HANG7 dataset ***Wang et al.*** (***2023***) for type-1 narcolepsy (T1N) versus other sleep disorders.

| Dataset | Method | BACC | SENS | SPEC |
|---|---|---|---|---|
| **MNC (CV)** | Hypnogram | $0.843_{(0.797–0.885)}$ | $0.828_{(0.738–0.907)}$ | $0.858_{(0.819–0.894)}$ |
| | Hypnodensity | $0.851_{(0.805–0.894)}$ | $0.816_{(0.729–0.896)}$ | $0.886_{(0.854–0.917)}$ |
| | BaseNet | $0.797_{(0.745–0.850)}$ | $0.671_{(0.574–0.775)}$ | $0.924_{(0.895–0.951)}$ |
| | From scratch | $0.839_{(0.787–0.888)}$ | $0.754_{(0.656–0.848)}$ | $0.923_{(0.896–0.950)}$ |
| | Pretrained | $\mathbf{0.929}_{(0.896–0.960)}$ | $\mathbf{0.915}_{(0.854–0.971)}$ | $\mathbf{0.944}_{(0.917–0.967)}$ |
| **MNC → HANG7** | Hypnogram | $0.724_{(0.575–0.874)}$ | $0.616_{(0.318–0.900)}$ | $0.832_{(0.739–0.915)}$ |
| | Hypnodensity | $0.755_{(0.605–0.896)}$ | $0.692_{(0.400–0.933)}$ | $0.818_{(0.729–0.908)}$ |
| | BaseNet | $0.693_{(0.549–0.831)}$ | $0.695_{(0.429–0.933)}$ | $0.690_{(0.579–0.797)}$ |
| | From scratch | $0.753_{(0.615–0.873)}$ | $0.773_{(0.500–1.000)}$ | $0.733_{(0.623–0.831)}$ |
| | Pretrained | $\mathbf{0.840}_{(0.733–0.917)}$ | $\mathbf{0.922}_{(0.727–1.000)}$ | $0.759_{(0.652–0.851)}$ |

Values are reported as point estimates with 95% confidence intervals (shown as subscripts). Confidence intervals were estimated using bootstrap resampling. BACC, balanced accuracy; SENS, sensitivity; SPEC, specificity. Hypnodensity denotes an empirical feature-based XGBoost classifier using hypnodensity features derived from sleep stage probability distributions. Bold values indicate the best performance for each dataset.

#### Supplementary figures

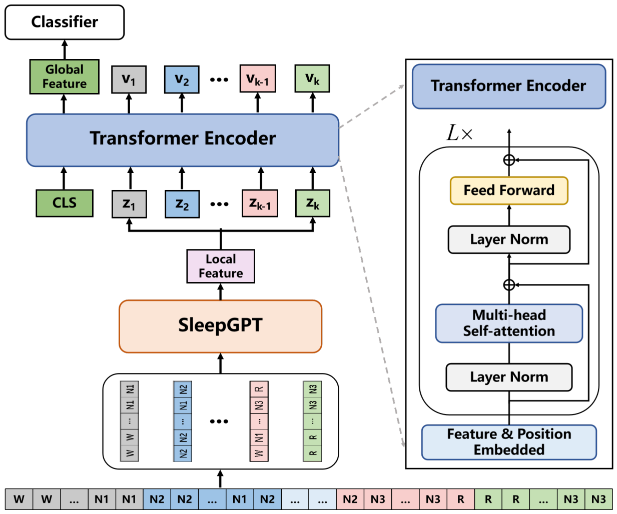

**Figure S1.** The hierarchical transformer network for stage sequence-based sleep disorder diagnosis. The HTN model comprises a local feature extractor, i.e., the SleepGPT model, a transformer encoder for global feature extraction, and a classification head to perform diagnosis. The configuration of the global transformer encoder is shown on the right. On the bottom is an example of a whole-night sleep stage sequence, which is partitioned into non-overlapping segments that are subsequently input into the HTN model.

#### References

**Chan W**, Jaitly N, Le Q, Vinyals O. Listen, attend and spell: A neural network for large vocabulary conversational speech recognition. In: *2016 IEEE International Conference on Acoustics, Speech and Signal Processing (ICASSP)*; 2016. p. 4960–4964. doi: 10.1109/ICASSP.2016.7472621.

**Dosovitskiy A**, Beyer L, Kolesnikov A, Weissenborn D, Zhai X, Unterthiner T, Dehghani M, Minderer M, Heigold G, Gelly S, Uszkoreit J, Houlsby N. An Image is Worth 16x16 Words: Transformers for Image Recognition at Scale. In: *International Conference on Learning Representations*; 2021. https://openreview.net/forum?id=YicbFdNTTy.

**Ghassemi MM**, Moody BE, Lehman LH, Song C, Li Q, Sun H, Mark RG, Westover MB, Clifford GD. You Snooze, You Win: the PhysioNet/Computing in Cardiology Challenge 2018, Journal Article. Computing in Cardiology (2010). 2018; 45. doi: 10.22489/cinc.2018.049.

**Goldberger AL**, Amaral LAN, Glass L, Hausdorff JM, Ivanov PC, Mark RG, Mietus JE, Moody GB, Peng CK, Stanley HE. PhysioBank, PhysioToolkit, and PhysioNet. Circulation. 2000; 101(23):e215–e220. https://www.ahajournals.org/doi/abs/10.1161/01.CIR.101.23.e215, doi: 10.1161/01.CIR.101.23.e215.

**Iber C**, Ancoli-Israel S, Chesson AL, Quan S. The AASM Manual for the Scoring of Sleep and Associated Events: Rules, Terminology and Technical Specifications. American Academy of Sleep Medicine. 2007; .

**Karita S**, Soplin N, Watanabe S, Delcroix M, Ogawa A, Nakatani T. Improving transformer-based end-to-end speech recognition with connectionist temporal classification and language model integration. Proceedings of the Annual Conference of the International Speech Communication Association, INTERSPEECH. 2019; 2019-September:1408–1412. doi: 10.21437/Interspeech.2019-1938, publisher Copyright: Copyright © 2019 ISCA; 20th Annual Conference of the International Speech Communication Association: Crossroads of Speech and Language, INTERSPEECH 2019 ; Conference date: 15-09-2019 Through 19-09-2019.

**Kemp B**, Zwinderman AH, Tuk B, Kamphuisen HAC, Oberye JJL. Analysis of a sleep-dependent neuronal feedback loop: the slow-wave microcontinuity of the EEG. IEEE Transactions on Biomedical Engineering. 2000; 47(9):1185–1194. doi: 10.1109/10.867928.

**Kenton JDMWC**, Toutanova LK. BERT: Pre-training of Deep Bidirectional Transformers for Language Understanding. In: *Proceedings of NAACL-HLT*; 2019. p. 4171–4186.

**Khalighi S**, Sousa T, Santos JM, Nunes U. ISRUC-Sleep: A comprehensive public dataset for sleep researchers. Computer Methods and Programs in Biomedicine. 2016; 124:180–192. https://www.sciencedirect.com/science/article/pii/S0169260715002734, doi: https://doi.org/10.1016/j.cmpb.2015.10.013.

**Lopez-Larraz E**, Sierra-Torralba M, Clemente S, Fierro G, Oriol D, Minguez J, Montesano L, Klinzing JG. Bitbrain Open Access Sleep Dataset. . 2024; doi: doi:10.18112/openneuro.ds005555.v1.0.0.

**Mignot E**, Lammers GJ, Ripley B, Okun M, Nevsimalova S, Overeem S, Vankova J, Black J, Harsh J, Bassetti C, Schrader H, Nishino S. The Role of Cerebrospinal Fluid Hypocretin Measurement in the Diagnosis of Narcolepsy and Other Hypersomnias. Archives of Neurology. 2002; 59(10):1553–1562. https://doi.org/10.1001/archneur.59.10.1553, doi: 10.1001/archneur.59.10.1553.

**Mousavi S**, Afghah F, Acharya UR. SleepEEGNet: Automated sleep stage scoring with sequence to sequence deep learning approach, Journal Article. PLoS One. 2019; 14(5):e0216456. https://www.ncbi.nlm.nih.gov/pubmed/31063501, doi: 10.1371/journal.pone.0216456.

**O'Reilly C**, Gosselin N, Carrier J, Nielsen T. Montreal Archive of Sleep Studies: an open-access resource for instrument benchmarking and exploratory research. Journal of Sleep Research. 2014; 23(6):628–635. https://onlinelibrary.wiley.com/doi/abs/10.1111/jsr.12169, doi: https://doi.org/10.1111/jsr.12169.

**Pappagari R**, Zelasko P, Villalba J, Carmiel Y, Dehak N. Hierarchical Transformers for Long Document Classification. In: *2019 IEEE Automatic Speech Recognition and Understanding Workshop (ASRU)*; 2019. p. 838–844. doi: 10.1109/ASRU46091.2019.9003958.

**Phan H**, Andreotti F, Cooray N, Chen OY, De Vos M. Joint Classification and Prediction CNN Framework for Automatic Sleep Stage Classification, Journal Article. IEEE Transactions on Biomedical Engineering. 2019; 66(5):1285–1296. https://www.ncbi.nlm.nih.gov/pubmed/30346277, doi: 10.1109/TBME.2018.2872652.

**Phan H**, Andreotti F, Cooray N, Chen OY, De Vos M. SeqSleepNet: End-to-End Hierarchical Recurrent Neural Network for Sequence-to-Sequence Automatic Sleep Staging, Journal Article. IEEE Transactions on Neural Systems and Rehabilitation Engineering. 2019; 27(3):400–410. https://www.ncbi.nlm.nih.gov/pubmed/30716040, doi: 10.1109/TNSRE.2019.2896659.

**Phan H**, Chen OY, Tran MC, Koch P, Mertins A, De Vos M. XSleepNet: Multi-View Sequential Model for Automatic Sleep Staging, Journal Article. IEEE Transactions on Pattern Analysis and Machine Intelligence. 2022; 44(9):5903–5915. https://www.ncbi.nlm.nih.gov/pubmed/33788679, doi: 10.1109/TPAMI.2021.3070057.

**Quan SF**, Howard BV, Iber C, Kiley JP, Nieto FJ, O'Connor GT, Rapoport DM, Redline S, Robbins J, Samet JM, Wahl PW. The Sleep Heart Health Study: Design, Rationale, and Methods. Sleep. 1997; 20(12):1077–1085. https://doi.org/10.1093/sleep/20.12.1077, doi: 10.1093/sleep/20.12.1077.

**Radford A**, Narasimhan K, Salimans T, Sutskever I. Improving language understanding with unsupervised learning. . 2018; .

**Radford A**, Wu J, Child R, Luan D, Amodei D, Sutskever I, et al. Language models are unsupervised multitask learners. OpenAI blog. 2019; 1(8):9.

**Rao K**, Sak H, Prabhavalkar R. Exploring architectures, data and units for streaming end-to-end speech recognition with RNN-transducer. In: *2017 IEEE Automatic Speech Recognition and Understanding Workshop (ASRU)*; 2017. p. 193–199. doi: 10.1109/ASRU.2017.8268935.

**Stephansen JB**, Olesen AN, Olsen M, Ambati A, Leary EB, Moore HE, Carrillo O, Lin L, Han F, Yan H, Sun YL, Dauvilliers Y, Scholz S, Barateau L, Hogl B, Stefani A, Hong SC, Kim TW, Pizza F, Plazzi G, et al. Neural network analysis of sleep stages enables efficient diagnosis of narcolepsy, Journal Article. Nature Communications. 2018; 9(1):5229. https://www.ncbi.nlm.nih.gov/pubmed/30523329, doi: 10.1038/s41467-018-07229-3.

**Supratak A**, Dong H, Wu C, Guo Y. DeepSleepNet: A Model for Automatic Sleep Stage Scoring Based on Raw Single-Channel EEG, Journal Article. IEEE Transactions on Neural Systems and Rehabilitation Engineering. 2017; 25(11):1998–2008. https://www.ncbi.nlm.nih.gov/pubmed/28678710, doi: 10.1109/TNSRE.2017.2721116.

**Supratak A**, Guo Y. TinySleepNet: An Efficient Deep Learning Model for Sleep Stage Scoring based on Raw Single-Channel EEG. In: *The 42nd Annual International Conference of the IEEE Engineering in Medicine & Biology Society (EMBC)*; 2020. p. 641–644. doi: 10.1109/EMBC44109.2020.9176741.

**Terzano MG**, Parrino L, Sherieri A, Chervin R, Chokroverty S, Guilleminault C, Hirshkowitz M, Mahowald M, Moldofsky H, Rosa A, Thomas R, Walters A. Atlas, rules, and recording techniques for the scoring of cyclic alternating pattern (CAP) in human sleep. Sleep Medicine. 2001; 2(6):537–553. https://www.sciencedirect.com/science/article/pii/S1389945701001496, doi: https://doi.org/10.1016/S1389-9457(01)00149-6.

**Vaswani A**, Shazeer N, Parmar N, Uszkoreit J, Jones L, Gomez AN, Kaiser L, Polosukhin I. Attention is All you Need. In: *Advances in Neural Information Processing Systems*, vol. 30; 2017. .

**Wang J**, Zhao S, Zhou Y, Jiang H, Yu Z, Li T, Li S, Pan G. Narcolepsy Diagnosis With Sleep Stage Features Using PSG Recordings. IEEE Transactions on Neural Systems and Rehabilitation Engineering. 2023; 31:3619–3629. doi: 10.1109/TNSRE.2023.3312396.

**Wolpert EA**. A Manual of Standardized Terminology, Techniques and Scoring System for Sleep Stages of Human Subjects. Archives of General Psychiatry. 1969; 20(2):246–247. https://doi.org/10.1001/archpsyc.1969.01740140118016, doi: 10.1001/archpsyc.1969.01740140118016.

**Yang Z**, Yang D, Dyer C, He X, Smola A, Hovy E. Hierarchical Attention Networks for Document Classification. In: *Proceedings of the 2016 Conference of the North American Chapter of the Association for Computational Linguistics: Human Language Technologies* San Diego, California: Association for Computational Linguistics; 2016. p. 1480–1489. https://aclanthology.org/N16-1174, doi: 10.18653/v1/N16-1174.

**Zhang GQ**, Cui L, Mueller R, Tao S, Kim M, Rueschman M, Mariani S, Mobley D, Redline S. The National Sleep Research Resource: towards a sleep data commons. Journal of the American Medical Informatics Association. 2018; 25(10):1351–1358. https://doi.org/10.1093/jamia/ocy064, doi: 10.1093/jamia/ocy064.
